# Projections of the incidence of COVID-19 in Japan and the potential impact of a Fall 2023 COVID-19 Vaccine

**DOI:** 10.1101/2023.10.24.23297475

**Authors:** M Kohli, M Maschio, A Lee, A Igarashi

## Abstract

**Background:** The study objective was to estimate the incidence of COVID-19 infection, hospitalization, and deaths in Japan from September 2023 to August 2024 and potential impact of a Fall 2023 COVID-19 vaccine for adults ≥18 years on these outcomes.

**Methods:** A previously developed Susceptible-Exposed-Infected-Recovered model for the United States (US) was adapted to Japan. The numbers of symptomatic infections, COVID-19– related hospitalizations, and deaths were calculated. Given differences in vaccination coverage, masking practices and social mixing patterns between the US and Japan, all inputs were updated to reflect the Japanese context. Vaccine effectiveness (VE) values are hypothetical, but predicted based on existing VE values of bivalent BA.4/BA.5 boosters against BA.4/BA.5 in Japan, from the VERSUS test-negative case-control study. Sensitivity analyses were performed.

**Results:** The base case model predicts overall that there will be approximately 35.2 million symptomatic COVID-19 infections, 690,000 hospitalizations, and 62,000 deaths in Japan between September 2023 and August 2024. If an updated COVID-19 vaccine is offered to all adults aged 18 years and older in Fall 2023, the model predicts that 7.3 million infections, 275,000 hospitalizations and 26,000 deaths will be prevented. If vaccines are only given to those aged 65 years and older, only 2.9 million infections, 180,000 hospitalizations and 19,000 deaths will be prevented. Sensitivity analysis results suggest that hospitalizations and deaths prevented are most sensitive to initial vaccine effectiveness (VE) against infection and hospitalizations, and the waning rate associated with VE against infection. Symptomatic infections prevented was most sensitive to initial VE against infection and VE waning.

**Conclusions:** Results suggest that a Fall 2023 COVID-19 vaccine would reduce total numbers of COVID-19 related infections, hospitalizations, and deaths.

## Introduction

While the COVID-19 vaccination program in Japan started more slowly than in other countries such as the United States (US), by the end of 2021 approximately 80% of the Japanese population had received two doses of a COVID-19 vaccine.^1^ Vaccine uptake was highest in the older age groups with coverage in those ages 65 years and older estimated at 92.6%.^2^ Since completion of the initial campaign, the Japanese Ministry of Health provided additional monovalent booster doses in Spring 2022, with a similarly high uptake in the older age groups.^3,4^ Additional monovalent doses and a bivalent COVID-19 vaccine have also been offered to those at highest risk, including those ages 65 years and older. The Japanese Ministry of Health has recently stated that all individuals ages 6 months and older are eligible to receive a government-funded updated monovalent vaccine in Fall 2023.^5^ Furthermore, they have recommended that those considered high-risk for severe outcomes, including those ages 65 years and older and those ages 5 to 64 years with immunocompromising conditions, receive a booster.

Japan has one of the oldest populations in the world and yet experienced one of the lowest rates of COVID-19 mortality during the pandemic. In 2021, the median age of the Japanese population was 48.4 years, whereas it was only 37.7 years in the US.^6^ Moreover, in 2023, 29.2% of the Japanese population was estimated to be 65 years or over,^7^ compared to only 18.1% in the US. Despite this difference in the age structure of these two countries, the COVID-19 mortality rate for 2020-21 in Japan was reported to be only 7.3 per 100,000, while it was 130.6 per 100,000 in the United States.^8^ Similarly, the estimated the excess mortality rate in Japan during this time was 44.1 per 100,000 compared to 179.3 per 100,000 in the US.^8^

Overall, the dynamics of the COVID-19 pandemic were quite different between Japan and the US. The Institute for Health Metrics and Evaluation (IHME) has estimated that the highest peak of COVID-19 infections to date in Japan occurred between July and October 2022.^9^ This contrasts with the US, where the highest peak occurred between December 2021 and January 2022 with the emergence of Omicron.^10^ The number of daily deaths in Japan has increased in 2022 and 2023, while on average, the numbers have been decreasing in the US. Indeed, the Japanese Ministry of Health has warned that the rate of hospitalization and severe disease related to COVID-19 infections is still high, particularly in older adults. Restrictions to encourage social distancing measures were only officially removed in Japan in May 2023 and mobility has not yet returned to pre-pandemic levels. ^11,12^ In addition, despite the lifting of masking mandates, 55% of people indicate that they plan to continue to wear masks all or some of the time.^13^ As these patterns of social interactions shift, the epidemiology of COVID-19 will also continue to evolve.

We have previously developed a Susceptible-Exposed-Infected-Recovered (SEIR) model to project the future incidence of COVID-19 infections in the US and determine the potential future impact of COVID-19 vaccines. In this manuscript, we describe the adaptation of that model to the Japanese context in order to predict the future burden of COVID-19 infections in Japan. The specific objectives of this analysis were to estimate the incidence of COVID-19 infection, hospitalization, and deaths in Japan from September 2023 to August 2024 and to estimate the potential impact of a Fall 2023 COVID-19 vaccine on these outcomes.

## Methods

### Model Structure: Simulation of Total Infections

The number of total COVID-19 infections (both asymptomatic and symptomatic) is estimated using a Susceptible-Exposed-Infected-Recovered (SEIR) model that was created to project the incidence of COVID-19 in the US in Fall 2022^14^ and updated to conduct projections for Fall 2023^15^. Briefly, individuals in the Susceptible (S) compartment are subject to a rate of infection that is updated in each model time step, or 1 day period, during the simulation. Infection incidence is dependent on the number of susceptible people in the population as well as the rate of effective contacts between susceptible and infected individuals. Effective contacts are a function of the rate of contact and the transmissibility of the virus per contact. With an infected contact, individuals move to the Exposed (E) compartment, which represents a latent or pre-infectious state, and then to the Infected (I) compartment. Those in the I compartment can transmit the infection but may be symptomatic or asymptomatic. When the infection is cleared, individuals move to the Recovered (R) compartment where they are immune to another infection. As this acquired natural immunity fades, individuals return to the S compartment.

The model has six different vaccine strata that each contain SEIR states: unvaccinated; received primary series; received monovalent booster 1; received monovalent booster 2; received bivalent booster, and received Fall 2023 vaccine. Vaccination reduces the risk of an effective contact and a different average vaccine effectiveness (VE) is calculated for each model stratum. VE is a function of time since vaccination and the match of the vaccine to the current circulating strain. Individuals in the S or R states may receive a new vaccination which causes them to transition to the parallel S or R states in the appropriate vaccine stratum. The differential equations for this model are presented in the Technical Appendix. To reflect difference in implementation of the COVID-19 vaccination programs, the Japanese model differs from the US model structure in the allowed transitions between the vaccine strata; these are also described in the Technical Appendix.

### Model Inputs: SEIR Model

Given the differences in vaccination coverage, masking practices and social mixing patterns between the US and Japan, all inputs were updated to reflect the Japanese context. All inputs are described in the Technical Appendix. As with the US model, the simulation started in January 2020. The period between January 2020 and August 31, 2023 was used as a “calibration period” or a “burn-in” period. The model was run during this period in order to calculate the residual VE in each vaccine stratum on August 31, 2023. In a calibration process, also described in the Technical Appendix, the transmissibility parameter was varied so that the model reproduced an estimated target of infections throughout this timeframe so that on August 31, 2023, a reasonable proportion of the cohort were in the R state and had natural immunity.

For the base case model, the target from June 2022 onwards was estimated using infection data from the Tokyo precinct.^16^ This infection data was extrapolated to the entire Japanese population, then doubled assuming that only 50% of symptomatic infections are detected and come to medical attention, then adjusted to include both symptomatic and asymptomatic infections.

For scenario analyses, alternative calibration targets were created in order to test the impact of assumptions made for the base case. To vary the level of natural immunity in the population, a different calibration target was developed. Instead of doubling the number of observed infections as in the base case, the number was multiplied by 2.5 and 1.5. In addition, a calibration using the base case targets, but a higher monthly waning rate against infections for the vaccines during the Omicron period (6.58%) was conducted. This last calibration was expected to produce a different level of both residual VE and natural immunity at the start of the analytic period on September 1, 2023. The results of the various calibrations are presented in the Technical Appendix.

The analysis timeframe ran from September 1, 2023 to August 31, 2024. During this time period, the transmissibility parameter was held constant. A scenario where there were no further vaccinations after August 31, 2023 was then compared to a scenario where a new vaccine was administered in Fall 2023 in order to determine the impact of the vaccine on the number of infections.

For scenario analyses, inputs for the projection period between September 1, 2023 and August 31, 2024 were varied. The characteristics of the sine curve representing seasonality were modified in order to change the mobility scalar and the associated contact patterns throughout the year. Figure 11 in Technical Appendix contrasts the mobility scalar value for the base case and for this new scenario. In addition, the transmissibility parameter was varied from 0.32 in the base case to 0.25 and 0.40 in additional scenario analyses.

A summary of the different COVID-19 incidence projections for the base case and all scenario analyses, without a Fall 2023 vaccination, is presented in the Technical Appendix (Figure 12).

### Fall 2023 COVID-19 Vaccine

While the Japanese Ministry of Health has deemed all individuals ages 6 months and older to be eligible for a Fall 2023 vaccine, this analysis focuses on the adult population only. For the base case analysis, it was assumed that a COVID-19 mRNA vaccine updated for Fall 2023 was administered to adults ages 18 years and older. In a scenario analysis, the vaccine was only administered to adults ages 65 years and older.

### Model Inputs: Fall 2023 COVID-19 Vaccine Coverage

The Fall 2023 COVID-19 vaccine campaign is assumed to be administered between September and December 2023 as shown in the Technical Appendix Figure 9. The maximum vaccine coverage for each age group was assumed to be the same as the maximum uptake rate observed for the first COVID-19 booster (Dose 3) administered in Japan.^3,4^ Table 1 displays the values used for base case and the sensitivity analyses.

**Table 1:**
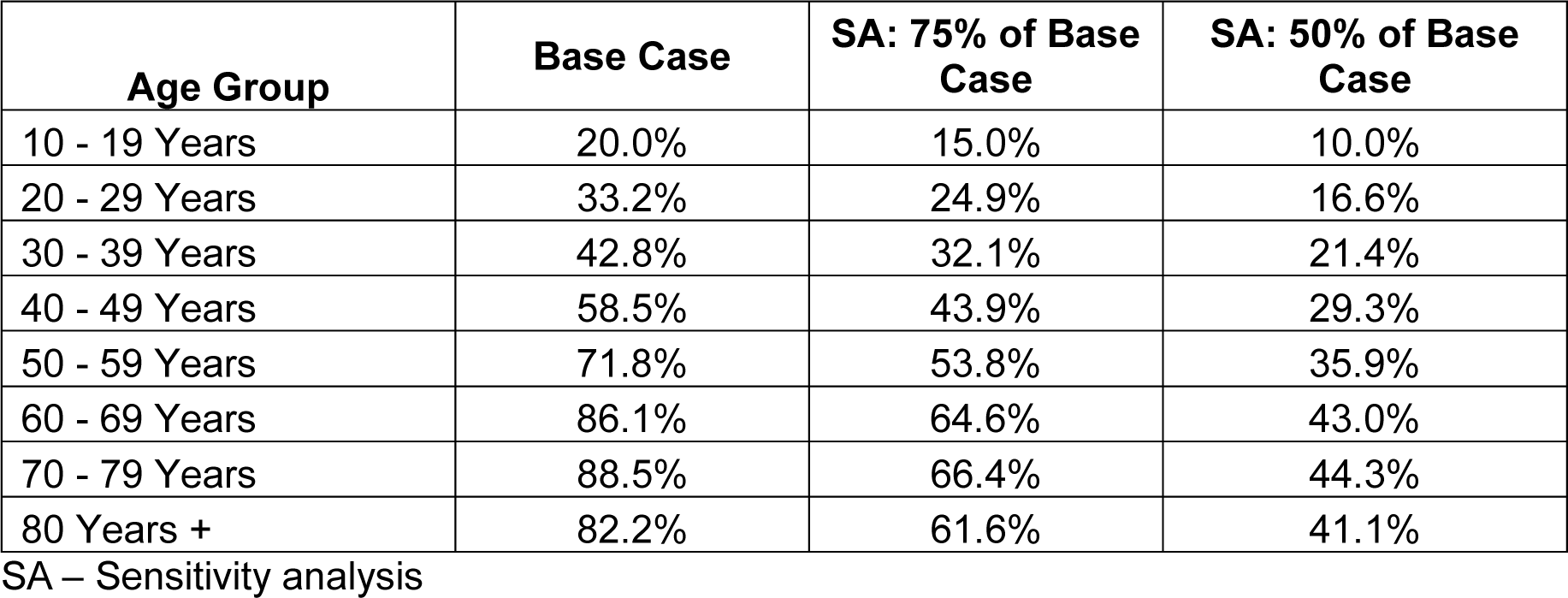
Maximum percent of population assumed to receive the Fall 2023 COVID-19 vaccine, by age group.

### Model Inputs: Fall 2023 Vaccine Effectiveness

The VE of the updated Fall 2023 vaccine is assumed to be well-matched to the circulating variant at the time. At time of writing, real-world VE data do not exist, as the model is projecting in the future. Therefore, VE values are hypothetical, but predicted based on existing VE values of bivalent BA.4/BA.5 boosters against BA.4/BA.5 in Japan, using data from the VERSUS study, a test-negative multi-centered case control study to estimate the real-world VE values.^17^ This study found the VE against infection and hospitalization in those ages 16-64 years in Japan to be 54.7% and 84.9%, respectively between October 1, 2022 and February 28, 2023.^18^ Thus, the model base case assumes VEs of 55% and 85% against infection and hospitalization (i.e. severe disease) respectively. Sensitivity analyses are included using an absolute ± 15% for infection (40-70%) and ±10% for hospitalization (75-95%). These values approximated the lower and upper 95% confidence intervals values observed in the VERSUS study when considering monovalent boosters against BA.1/BA.2 and bivalent boosters against BA.4/BA.5.^19–21^

The monthly decline in VE (waning rate) of the updated Fall 2023 was assumed to be 4.8% and 1.4% against infection and hospitalization, respectively. This was based on data from a meta-analysis by Higdon et al. (2022),^22^ which examined the duration of protection from monovalent vaccines against BA.1/BA.2. Sensitivity analyses were based on the 95% confidence intervals from this study: 3.1%-6.8% for infection and 0.6%-2.4% for hospitalization.

For the base case analysis, it was assumed that both natural immunity and the Fall 2023 vaccine was well-matched to the circulating variants. Two scenarios were conducted where a new variant with immune escape emerged in April 2024 and then in June 2024. VE and natural immunity were both assumed to drop immediately by 10% when this occurred.

Figure 2 presents the VE values used over time for the base case and sensitivity analyses for both infection and hospitalization outcomes.

**Figure 2:**
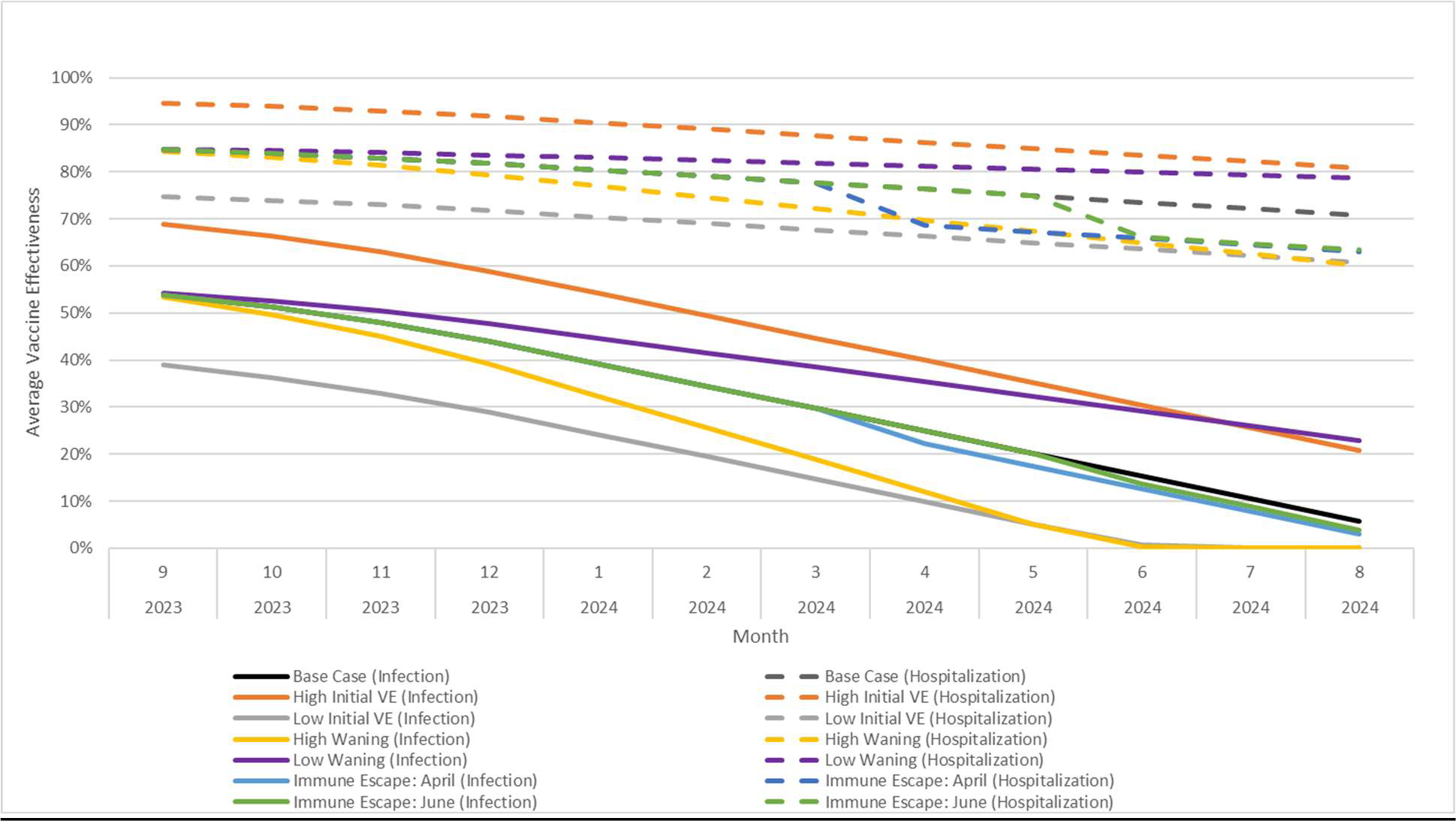
The estimated vaccine effectiveness of a COVID-19 vaccine from September 2023 to August 2024 (base case and scenario analyses).

### Model Structure: Simulation of Clinical Outcomes

The number of infections predicted by the SEIR model are summarized by month and output to a separate decision-tree model. First, the number of infections is reduced so that only the symptomatic infections enter this decision tree. All symptomatic cases have a probability of becoming severe enough to require hospitalization. The SEIR model simulation calculates the daily VE against infection for each vaccination stratum and uses these values to calculate daily incidence of infection. The model also calculates the incremental VE against hospitalization, which is averaged by month and then used by the decision tree to reduce the probability of hospitalization given symptomatic COVID-19 infection for people in each vaccination stratum. Finally, in this analysis, those who are hospitalized face a probability of death due to their COVID-19 infection.

### Model Inputs: Hospitalization and Mortality Rates

The overall rate of hospitalization amongst those who seek care for symptomatic COVID-19 infections was estimated based on the number of hospitalizations seen in Japan according to the National Institute of Infectious Disease (NIID) database from April 2022 to March 2023.^23^ The vaccination status of those hospitalized is not tracked, therefore the probability of hospitalization amongst those who are not vaccinated was estimated using assumptions about the level of residual VE in the population. (See the Technical Appendix for further details.) As the amount of residual VE is uncertain, two scenario analyses were created as shown in Table 2.

**Table 2:** Hospitalization and hospital mortality rates, by age group.

| Model Age Groups | Hospitalization amongst those with symptomatic infection (Unvaccinated population)* |  |  | Hospital Mortality Rates |
| --- | --- | --- | --- | --- |
|  | Base Case | SA: Increased hospitalization rate | SA: Decreased hospitalization rate |  |
| 0-11 years | 0.46% | 0.46% | 0.46% | 0.14% |
| 12-17 years | 0.18% | 0.19% | 0.16% | 0.14% |
| 18-29 years | 0.30% | 0.34% | 0.27% | 0.15% |
| 30-39 years | 0.47% | 0.54% | 0.41% | 0.15% |
| 40-49 years | 0.51% | 0.59% | 0.45% | 1.10% |
| 50-59 years | 1.04% | 1.21% | 0.91% | 1.10% |
| 60-64 years | 2.93% | 3.45% | 2.55% | 4.98% |
| 65-74 years | 8.62% | 10.13% | 7.51% | 7.07% |
| 75+ years | 13.76% | 15.86% | 12.15% | 12.91% |

\* Probability of hospitalization given a person with a symptomatic infection who seeks medical care.
SA – Scenario Analysis

The hospital mortality, also shown in Table 2, was calculated using two commercial databases provided by JAST Inc and DeSC Healthcare Inc.^24,25^ The JAST database is based on an employee’s insurance system. The DeSC database derives information from an insurance system for retired persons and one designed for persons aged 75 years and above in Japan. The JAST data was used to derive mortality rates for those below 60 years of age, while the DeSC data was used for those 60 years and older.

## Results

The base case results are summarized in Table 3. The base case model predicts overall that there will be approximately 35.2 million symptomatic COVID-19 infections, 690,000 hospitalizations, and 62,000 deaths in Japan between September 2023 and August 2024 if no COVID-19 vaccine is administered in this timeframe. If an updated COVID-19 vaccine is offered to all adults aged 18 years and older in Fall 2023, the model predicts that 72,542,457 will receive a vaccine and 7.3 million infections, 275,000 hospitalizations and 26,000 deaths will be prevented in the Japanese population.

**Table 3:** Base case analysis: Projected symptomatic COVID-19 infections and associated hospitalizations and deaths, with and without a COVID-19 updated Fall 2023 vaccine.

| Outcome | Vaccination: 18 years and older |  |  | Vaccination: 65 years and older |  |  |
| --- | --- | --- | --- | --- | --- | --- |
|  | No Vaccine | With Vaccine | Prevented (% decrease) | No Vaccine | With Vaccine | Prevented (% decrease) |
| Symptomatic Infection | 35,240,923 | 27,915,148 | 7,325,775 (21%) | 35,240,923 | 32,337,129 | 2,903,794 (8%) |
| Hospitalizations | 689,973 | 415,239 | 274,734 (40%) | 689,973 | 509,560 | 180,413 (26%) |
| Deaths | 61,738 | 35,881 | 25,857 (42%) | 61,738 | 42,266 | 19,472 (32%) |

The number of monthly infections and hospitalizations, by age group, over time is shown in Figure 3. Without vaccination, infection is predicted to be highest in the youngest and oldest age groups, while the majority of hospitalizations occur in those aged 65 and older. With a Fall 2023 vaccine administered to those ages 18 years and above, the number of infections decreases in all age groups, including in the younger age groups not eligible for the vaccine. As VE against infection declines to below 10% at the end of the year, the protective effect of the vaccine on the number of infections declines as well. Therefore, from May 2024 to August 2024, the model predicts a higher number of infections with the Fall 2023 vaccine than without the Fall 2023 vaccine. Over the entire year however, there are still predicted to be 21% fewer infections in the Japanese population. The number of hospitalizations in the Fall 2023 vaccine scenario is also higher than in the No Fall 2023 vaccine scenario between June 2024 to August 2024. The effect of the reduction in VE against infection is blunted because VE against hospitalization is more durable and is still above 70% by August 2024. Over the entire 1 year time horizon, the Fall 2023 vaccine prevents 40% of hospitalizations. As deaths are closely related to the number of hospitalizations, the vaccine is predicted to prevent 42% of COVID-19 attributable deaths.

**Figure 3:**
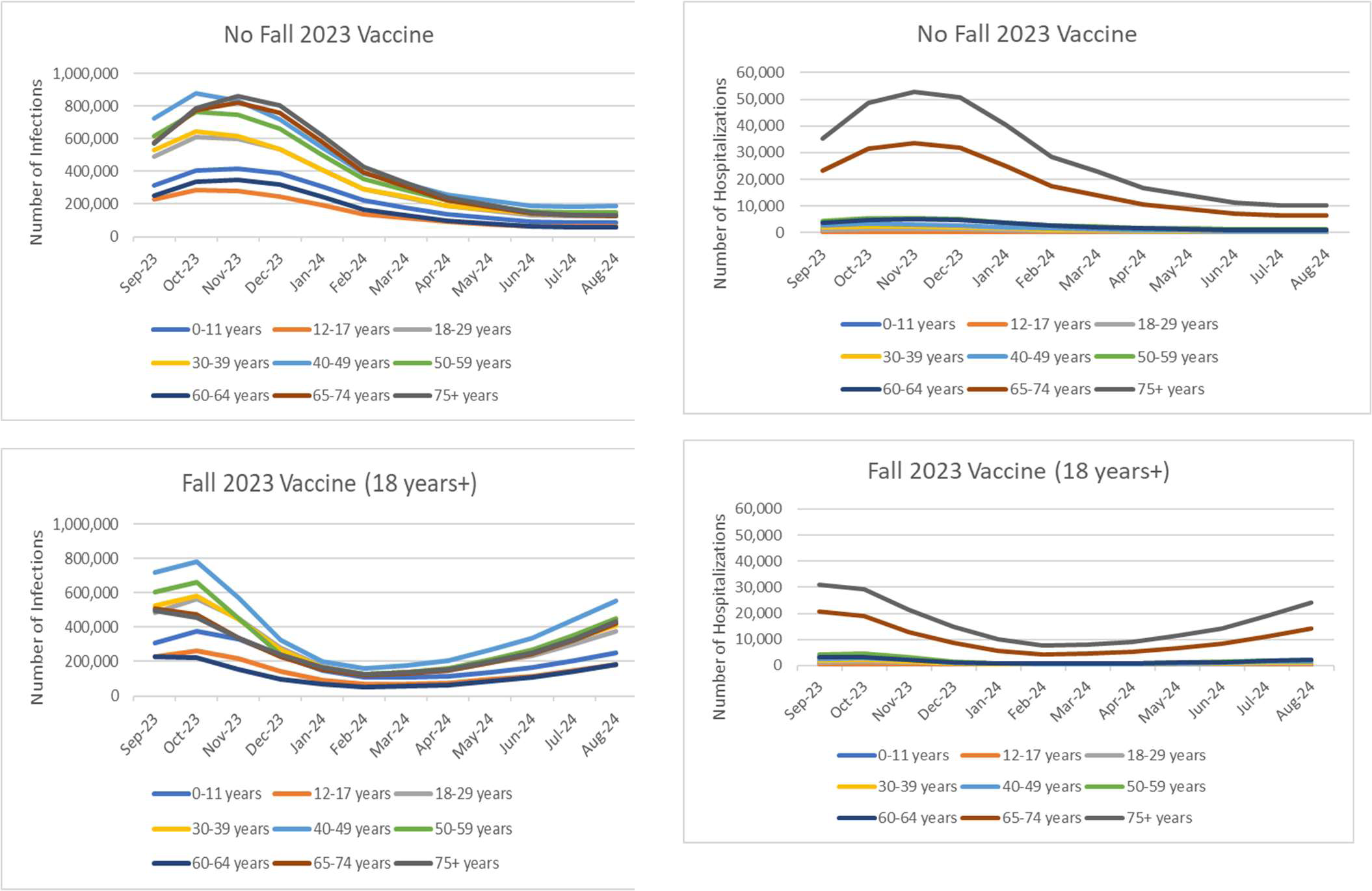
Projected number of monthly symptomatic infections and hospitalizations with and without a COVID-19 vaccine updated for Fall 2023, by age group.

If vaccines are only given to those aged 65 years and older, 32,116,154 will receive a vaccine and 2.9 million infections, 180,000 hospitalizations and 19,000 deaths will be prevented in the Japanese population. The number of vaccine doses administered to those aged 65 years and older is only 44% of the doses administered with the base case strategy of vaccinating the entire adult population. Therefore, fewer COVID-19 associated outcomes are also prevented: total infections in the Japanese population decrease by only 8%, hospitalizations by 26%, and deaths by 32%. As those 65 years and above are at highest risk of severe COVID-19 disease, vaccinating this age group has a greater impact as it prevents more hospitalizations and death per dose delivered.

As Figure 4 illustrates, in the base case, without vaccination, infection is predicted to peak in November 2023 and then decline gradually to a low in July and August 2024. If immune escape occurs, the loss in natural and residual vaccine immunity leads to an increase in infections in the months following occurrence of immune escape. If that occurs early enough in the year, as with the scenario with an immune escape variant in April 2024, a second peak of infection is expected to occur. With both immune escape scenarios, the overall number of infections is expected to be higher.

**Figure 4:**
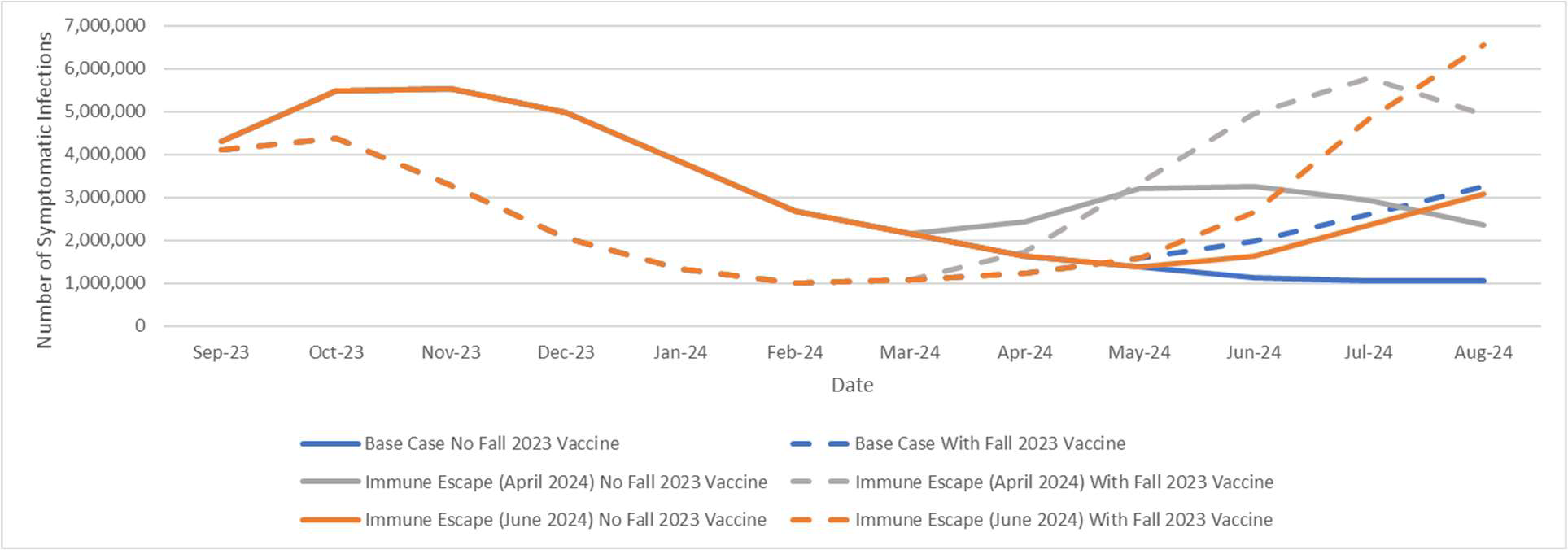
Projected COVID-19 incidence (symptomatic infection) with and without a COVID-19 vaccine updated for Fall 2023.

The results of the scenario analyses are shown in the tornado diagrams in Figure 5, which illustrate the variation in the predictions of the number of symptomatic infections, the number of hospitalizations and the number of deaths prevented. The results in the tornado diagram have been ordered from most to least impact on hospitalizations prevented. The importance of the different variables in driving the results is the same for both hospitalizations prevented and deaths prevented. The most important variables are the initial VE against infection and hospitalizations. This is followed by the waning rate associated with VE against infection. Next, vaccination coverage is important with lower coverage rates leading to a lower number of outcomes prevented. Finally, the number of hospitalizations prevented is impacted by the incidence rates, the hospitalization rates and the waning of hospitalization VE.

**Figure 5:**
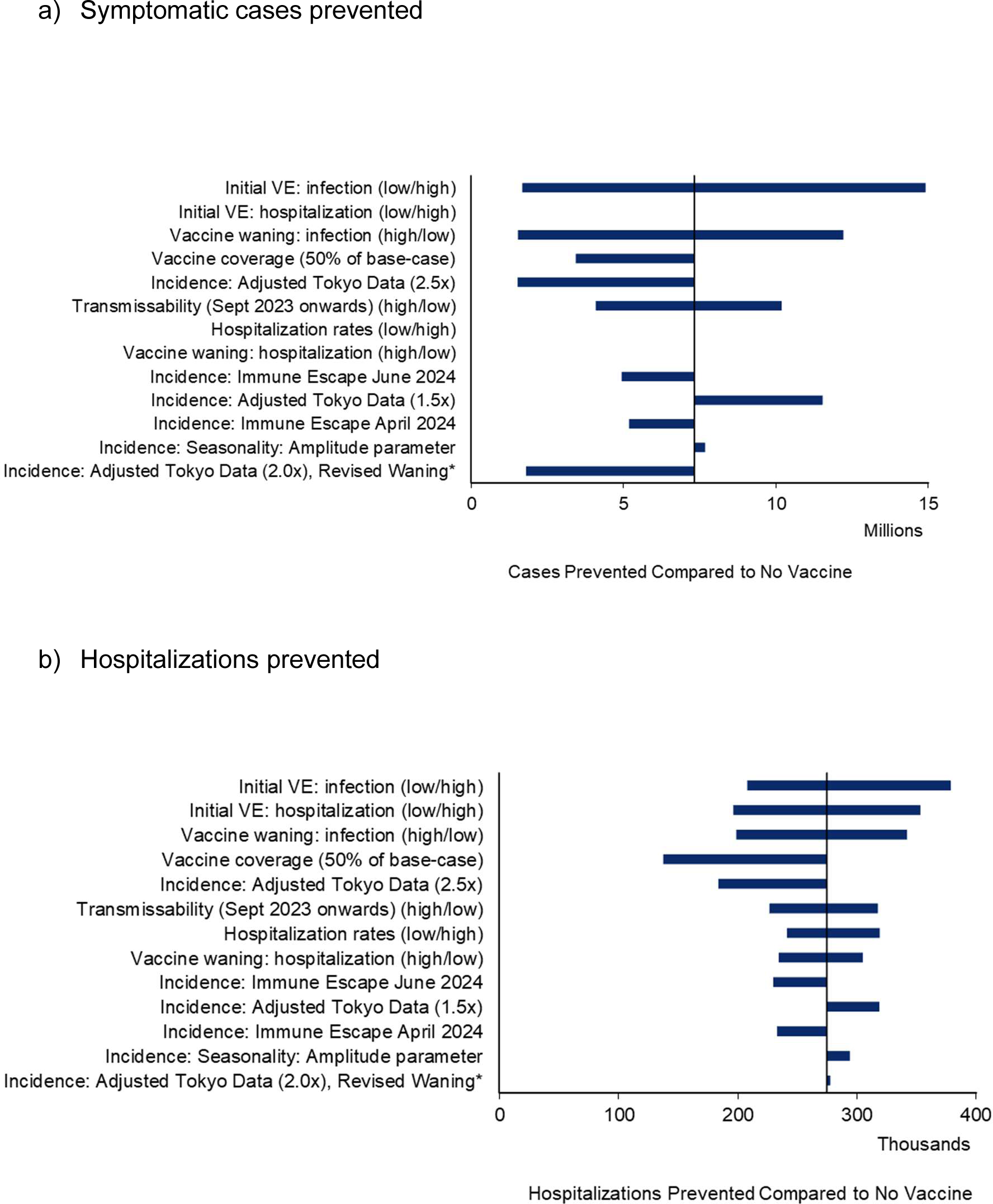
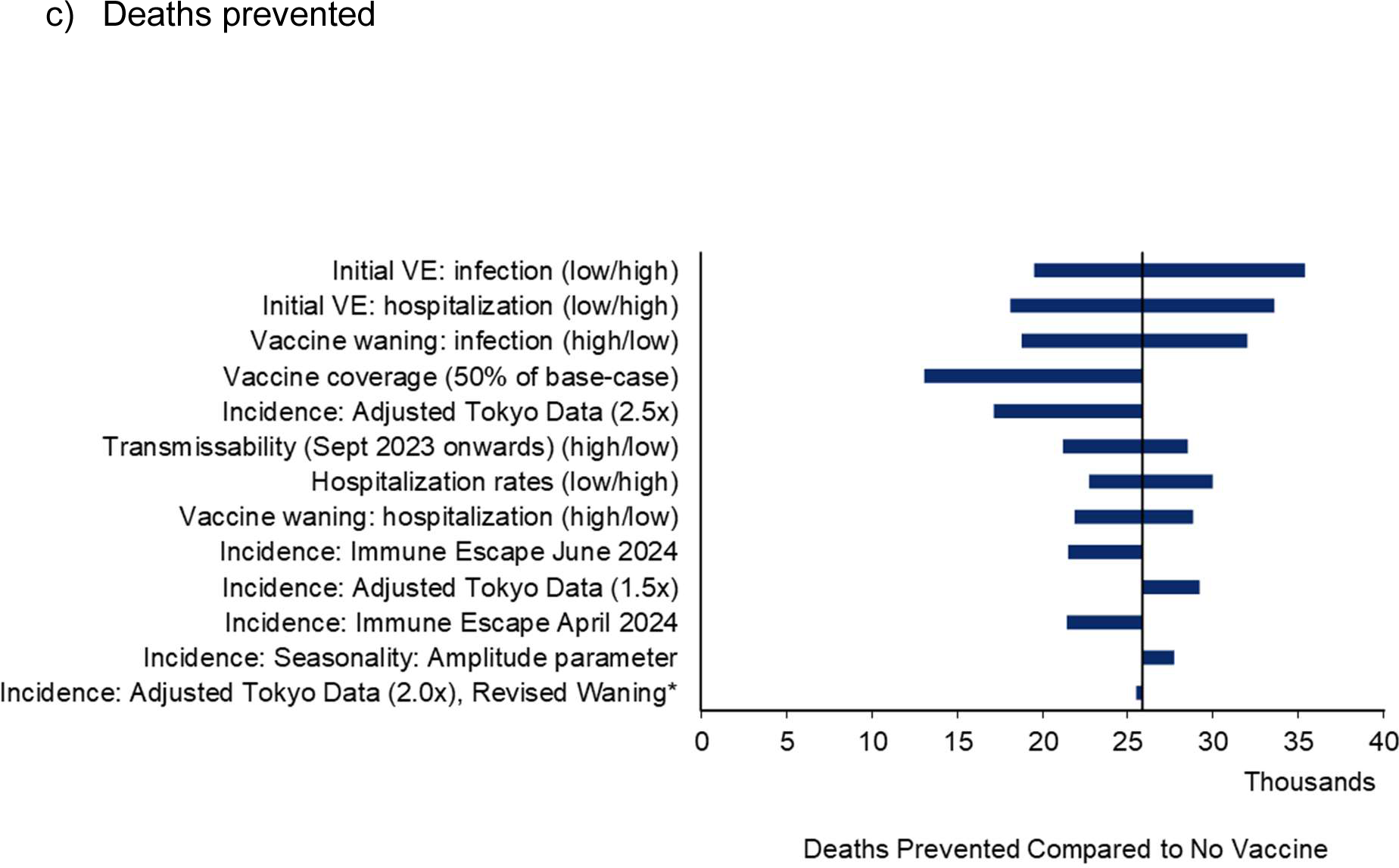
Impact of scenario analyses on the projected COVID-19 symptomatic infections, hospitalizations and deaths prevented with a COVID-19 vaccine updated for Fall 2023.

Considering the outcome of symptomatic infections, the initial VE against infection and VE waning are the most impactful variables. Next, the infections prevented are impacted by the change in incidence scenarios. Reducing the vaccine coverage reduces the infections prevented more than assuming that immune escape occurs in either April or June 2024.

To understand the impact of the different incidence scenarios, it is helpful to refer to additional data in the Technical Appendix. Figure 12 in the Technical Appendix displays the incidence of symptomatic infections by month when there is no fall 2023 vaccination campaign. Table 6 in the Technical Appendix displays the transmissibility parameters associated with the analysis period for the different incidence scenarios. Recalibration or changing the model inputs for the analysis time period will change the total number of infections predicted without a fall 2023 vaccination campaign by -9% to 13% (Table 8; Technical Appendix). More importantly, the timing of when infections occur changes, and fewer cases are prevented when the vaccine is not timed appropriately with the projected incidence. For example, changing the seasonality or calibrating to a lower number of Omicron infections (Scenario: Adjusted Tokyo Data (1.5X)) delays the peak of the incidence curve from November to December 2023. Therefore, more vaccines are delivered before the peak infection rate and 5% to 57% more infections are prevented (Table 8; Technical Appendix). When the transmissibility parameter is lowered, without a Fall 2023 vaccine, infections remain low until December and then start to increase. With Fall 2023 vaccination, most doses are delivered before December and therefore the vaccination campaign is predicted to 39% more symptomatic infections than in the base case. For three incidence scenarios, the Fall 2023 vaccine is predicted to prevent far fewer infections. When the transmission parameter is increased (Scenario: Higher transmissibility), the number of historical infections is higher (Scenario: Adjusted Tokyo Data (2.5X)), or the VE (infection) monthly waning over time is higher (Scenario: Adjusted Tokyo Data (2.0x), Revised Waning), there are more cases in September 2023 and the next incidence peak is shifted to Spring 2024 (See Figure 12 in the Technical Appendix). Overall, a Fall 2023 vaccination campaign is predicted to prevent 44%, 75% and 79% fewer cases respectively in these scenarios (Table 8; Technical Appendix). Primarily, this occurs because the cases occurring in September are not prevented.

## Discussion

In this analysis, we have attempted to quantify the potential annual burden of COVID-19 and impact of the COVID-19 mRNA vaccines modified for Fall 2023. Vaccinating the Japanese population aged18 years and older is predicted to prevent 21% of infections, 40% of hospitalizations and 42% of deaths across the year. Overall, the VE against infection is lower and wanes more quickly than protection against hospitalizations attributable to COVID-19.

While the epidemiology of COVID-19 varies between Japan and the US, the main variables that affect the estimated magnitude of benefit are similar. The most important driver of the impact of COVID-19 vaccines is their VE, including initial VE against infection and hospitalization, as well as the duration of protection provided. Here we have used the bivalent vaccine data from the Japanese VERSUS study to estimate the base case initial VE of the Fall 2023 vaccine. This prospective test-negative case-control study recruited individuals visiting medical institutions, including hospitals or clinics, nation-wide.^17^ VE ranges were developed based on past VE data from this same study from monovalent vaccines used during the Omicron period. The ranges for our monthly waning values come from studies of the original monovalent vaccines during the Omicron period. The existing studies of the updated Fall 2023 monovalent vaccines include immunological outcomes only.^26^ The results of these studies demonstrate that these updated vaccines provide a stronger antibody response to circulating COVID-19 viruses than either the bivalent or previous monovalent version, suggesting higher VE and potentially longer duration of protection.^27^ However, studies with clinical outcomes are required to validate these conclusions and they will not be available ahead of Fall 2023 when the vaccine will be implemented.

The COVID-19 variant has continued to evolve and it is possible that a new variant of concern that shows evidence of immune escape will emerge in the next year. In our simulation, emergence of such a variant was assumed to decrease natural and vaccine-mediated immunity, leading to an increase in symptomatic infections whether or not a new vaccine is given in Fall 2023. We have attempted to quantify the potential impact of immune escape by using example simulations. However, it is not possible to predict when a variant with immune escape will actually emerge or the degree of impact on either natural or vaccine-mediated immunity this event would have. Our analysis simply demonstrates the potential surge in infections that could occur with immune escape.

There is a lot of uncertainty in the incidence of infection and we have attempted to calibrate our model to reflect past rates of infection. However, as infection reporting has declined world-wide with the emergence of Omicron, it is unclear how many of those with infections actually seek medical care and are therefore officially reported. The number of past infections impacts the degree of natural immunity in the population and our estimate of virus transmissibility moving forward. We have tested different variables and our scenario analyses demonstrate that these assumptions impact the timing infection peaks between September 2023 and August 2024. The timing of vaccination relative to the timing of the increase in infection is an important predictor of clinical impact. Clearly, vaccines that are administered shortly prior to the start of a new wave of COVID-19 infections will be more effective than those administered months before an infection increase as waning immunity will impact their overall protection. Data on the weekly population-based rates of COVID-19 associated hospitalizations from the COVID-NET system suggest that the burden of disease associated with COVID-19 is decreasing in the US.^28^ However, the future of COVID-19 infection in Japan is more unclear given that 2022 and 2023 saw higher morbidity and mortality than 2020 or 2021.^23^ Further research using locally collected health insurance claim data and epidemiological data, similar to the prospective VERSUS VE data, is warranted to understand the current patterns of COVID-19 infections and related outcomes in Japan.

The hospitalization rates that we estimated based on the NIID hospitalization data were similar to the hospitalization probabilities amongst those seeking treatment in the JAST and DeSC database. However, these probabilities represent the average probability in a highly vaccinated population and the vaccination status of those admitted to hospital is not known. For this modelling exercise, we needed to estimate the counter-factual of what hospitalization rates would be without vaccination. In Japan, more than 50% of the hospitalizations due to COVID-19 have been in those over the age of 70 years. Any under or over estimation of the hospitalization rates, particularly in this older age group, will change our burden predictions. We have done some sensitivity analyses varying these rates to demonstrate the potential impact of this input.

## Conclusions

While the future incidence of COVID-19 is highly uncertain, a COVID-19 vaccine updated for fall 2023 is expected to reduce the morbidity and mortality experienced in the next year. Additional collection of local longitudinal data is warranted to understand the emerging patterns of COVID-19 infections and associated outcomes in Japan.

## Supporting information

Technical Appendix

## Data Availability

Additional details of the methods or outcomes from the model in the present study are available upon reasonable request to the authors

