## Supplementary material for "Projections of the incidence of COVID-19 in Japan and the potential impact of a Fall 2023 COVID-19 Vaccine": Technical Appendix

### The clinical and economic impact of COVID-19 Fall 2023 vaccines in Japan

#### Technical Appendix

**Tables**

**Figures**

#### 1. MODEL STRUCTURE

As described in the main manuscript, the SEIR model was created by adapting a model developed to project COVID-19 infection in the United States (US). The main differences between the US and Japanese model structures are the transitions between the vaccine stratum. The transitions between the vaccination strata in the Japanese model are described in Figure 1.

**Figure 1. Overview of the allowed transitions between the vaccination strata in the SEIR model.**

The box below is a simplified depiction of the movement between the vaccination strata in the SEIR model for Japan. The SEIR model has six different vaccination strata that each contain SEIR states: unvaccinated (U); received primary series (V); received monovalent booster 1 (B1); received monovalent booster 2 (B2); received bivalent booster (BB) and received Fall 2023 vaccine (F23). The green arrows represent allowed transitions between the strata. In the model simulation for Japan, individuals are eligible to receive the Fall 2023 vaccine if they had received primary series plus at least one booster since the start of COVID-19 vaccination.

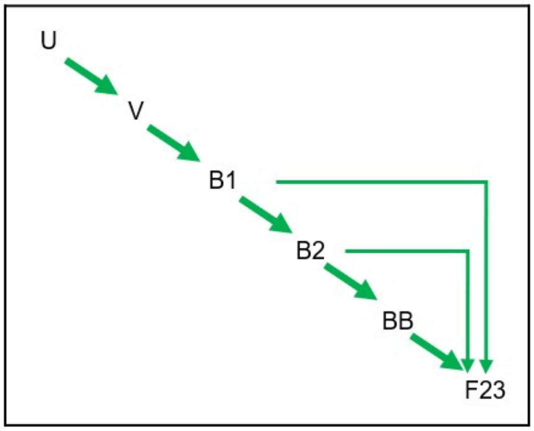

#### 2. SEIR MODEL INPUTS: BURN-IN PERIOD (JANUARY 2020 TO AUGUST 2023)

For the burn-in period (January 2020 to August 2023), the variables that determine the contact rate and the average vaccine effectiveness (VE) were estimated from data as described in Sections 2.2 - 2.3 below. The other model transitions (movements into the I, R, and S compartments) were estimated as described in Section 2.4. The transmissibility of the virus changed over time as new variants emerged. The transmissibility was therefore estimated through a calibration process described in Section 2.5. Any inputs that change in the analytic period are described in Section 3.

##### 2.1. Number of Susceptibles

All individuals in Japan were included in the model and were considered to be susceptible to COVID-19 infection at the start of the simulation (January 2020). The size of the Japanese population in 2020 by age was obtained from the United Nations Department of Economic and Social Affairs.[1]

**Table 1. The model population size**

| Age Group (Years) | Number |
|---|---|
| 0-9 | 9,525,018 |
| 10-19 | 11,103,195 |
| 20-29 | 12,131,456 |
| 30-39 | 13,694,755 |
| 40-49 | 18,060,661 |
| 50-59 | 16,333,785 |
| 60-69 | 15,466,156 |
| 70-79 | 16,582,824 |
| 80+ | 12,346,916 |
| **Total** | **125,244,766** |

##### 2.2. Number of Effective Contacts

The mixing patterns are based on contact matrices. As behaviors that impacted contact changed during the pandemic, the matrices are modified by a mobility index that accounts for social distancing and mask use.

###### 2.2.1. Mixing Patterns / Contact Matrices

Data on the age-specific mixing patterns in the general population for the US were obtained from Prem and colleagues.[2] In the published contact matrices, the population was partitioned into 5-year age bands, and all individuals aged 75 years and older were grouped together. For this analysis, the age-specific mixing patterns were first converted into 10-year age bands. Then, we assumed symmetry between the age groups (i.e., an effective contact between someone in age group *i* with someone from age group *j* is the same as an effective contact between someone in age group *j* with someone from age group *i),* weighted by the population estimates in age groups *i* and *j*.

Thus, $c_{ij} = \frac{1}{N_j} \times \frac{(c^*_{ij} \times N_j) + (c^*_{ji} \times N_i)}{2}$ , where:

$c_{ij}$ is the number of effective contacts between someone in age group *i* with someone from age group *j*

$c_{ij}^{*}$ is the number of effective contacts between someone in age group *i* with someone from age group *j* based on the original age-specific mixing patterns were first converted into 10-year age bands

$N_i$ is the population size in age group *i*.

The base contact matrix is presented in Table 2.

**Table 2. Base contact matrix used in the model before applying scaling factors**

| Age Group of Participant (Years) | Age Group of Contact (Years) | | | | | | | | |
|---|---|---|---|---|---|---|---|---|---|
| | **0-9** | **10-19** | **20-29** | **30-39** | **40-49** | **50-59** | **60-69** | **70-79** | **80+** |
| **0-9** | 4.059 | 0.763 | 0.468 | 1.354 | 0.568 | 0.444 | 0.386 | 0.224 | 0.290 |
| **10-19** | 0.890 | 8.066 | 1.152 | 1.153 | 1.312 | 0.835 | 0.360 | 0.343 | 0.455 |
| **20-29** | 0.596 | 1.259 | 4.633 | 2.278 | 1.587 | 1.321 | 0.474 | 0.216 | 0.243 |
| **30-39** | 1.947 | 1.422 | 2.571 | 4.976 | 2.566 | 1.738 | 1.045 | 0.502 | 0.571 |
| **40-49** | 1.076 | 2.133 | 2.363 | 3.384 | 3.912 | 2.335 | 0.982 | 0.828 | 0.956 |
| **50-59** | 0.761 | 1.228 | 1.778 | 2.073 | 2.112 | 2.969 | 1.133 | 0.612 | 0.785 |
| **60-69** | 0.627 | 0.501 | 0.605 | 1.180 | 0.841 | 1.073 | 2.681 | 1.191 | 0.921 |
| **70-79** | 0.390 | 0.512 | 0.296 | 0.608 | 0.761 | 0.622 | 1.277 | 2.011 | 1.882 |
| **80+** | 0.375 | 0.506 | 0.247 | 0.515 | 0.654 | 0.594 | 0.735 | 1.401 | 2.021 |

Note: Since the population size of the age groups are not of equal size, the transformed contact matrix is not strictly symmetric.

During the pandemic, the contact rate was reduced by behaviors such as social distancing and mask use. The magnitude of the impact is estimated as described in the next section.

###### 2.2.2. Social Distancing and Mask Use: Overall Scaling Factor

From January 30, 2020 onwards, it was assumed that regular patterns of interaction were modified because of social distancing and use of masks to reduce effective contacts between individuals. Data on social distancing patterns and mask use were used to adjust the base contact matrix based on these factors. Daily estimates of reduction in mobility and mask use were obtained from the Institute for Health Metrics and Evaluation (IHME) for the time period of

February 2020 through July 22, 2022, inclusive.[3] From August 23, 2022 to July 31, 2023, GPS data from Smartphones from the Tokyo station in Tokyo, Japan[4] were used to estimate the reduction in in mobility. From May 9, 2023 onwards mask usage was assumed to be 55% based on those who said they will continue to wear a mask or want to wear a mask, if situation allows in the LINE survey.[5] Between June 24, 2022 and May 9, 2023, a linear decrease in mask use over time was assumed. For the time period after July 31, 2023, a single seasonality parameter was also applied to simulate the normal seasonal change in mobility patterns.

Mask use represents the percentage of the population who say they always wear a mask in public. It was assumed that 100% mask usage is associated with a 30% reduction in transmission and that a reduction in mask usage impacts the reduction in transmission proportionately.[6] Therefore, the following scaling factor (due to mask use) was applied daily to the base contact matrix:

$$Scaling\ Factor_{Mask\ use} = 1 - (Mask\ use(\%) \times 0.30)$$

Daily estimates of the change in mobility were applied to reduce the number of contacts per person. Since age-specific data on changes in mobility were not available, the impact was applied equally to all age groups. Therefore, the following scaling factor (due to change in mobility) was applied daily to the base contact matrix:

$$Scaling\ Factor_{Mobility} = 1 + Change\ in\ mobility\ (\%)$$

After July 31, 2023, a standard sinusoidal function was applied to vary the rate of contact by season.[7] The assumed peak was February 15 to account for greater time spent indoors in the winter. The assumed trough was August 15 to account for greater time spent outdoors in summer. The trough and peak were 95% and 105% of normal contact respectively.[8]

We define the seasonality function over time applied to the transmissibility parameter that the transition rate is a function of time $\beta(t)$ as follows:

$$Seasonality = \left(1 + \frac{\phi}{2}\sin(2\pi t + \omega)\right)$$

where $\phi$ denotes the amplitude of seasonality (0.1 in the base case)

and $\omega$ is the phase shift of the sine function such that the peak was February 15 and the trough was August 15.

The overall scaling factor is thus calculated as:

| | | |
|---|---|---|
| Overall Scaling Factor | $[Scaling\ Factor_{Mask\ use}] \times [Scaling\ Factor_{Mobility}]$ | from January 30, 2020 to July 31, 2023; Constant after July 31, 2023 |
| | Seasonality estimate | after July 31, 2023 |

| | | |
|---|---|---|
| Overall Scaling Factor | $[1-(Mask\ use(\%)\times 0.30)] \times [1 + Change\ in\ mobility\ (\%)]$ | from January 30, 2020 to July 31, 2023; Constant after July 31, 2023 |
| | $\left(1+\frac{\phi}{2}\sin(2\pi t+\omega)\right)$ | after July 31, 2022 |

The final monthly pattern for the scaling factor for both the burn-in period (January 2020 – July 2023) and the analysis period (September 2023 – August 2024) is shown in Figure 2.

**Figure 2. Mobility scaling factor over time (burn-in and analysis period)**

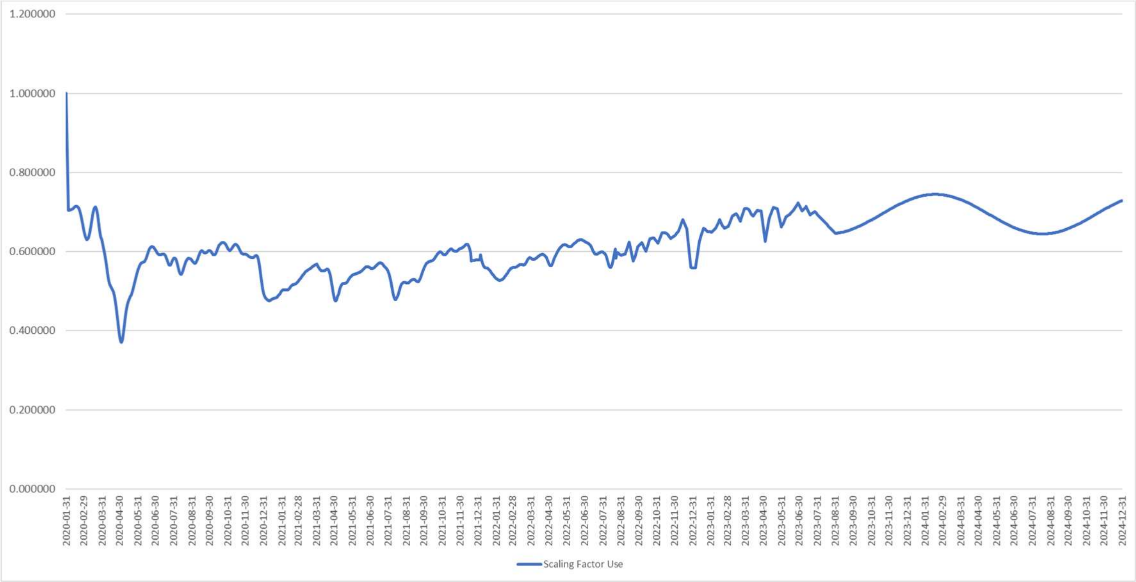

##### 2.3. Reduction in Effective Contacts Due to Vaccination

###### 2.3.1. Vaccine Coverage

A proportion of the population moves into the primary series and booster model strata according to vaccine uptake data from the Ministry of Health, Labour and Welfare of Japan website and the Official Website of the Prime Minister of Japan and His Cabinet as illustrated in Figure 3 to Figure 6.[9,10]

**Figure 3. Percent of the population who have completed their primary series, by age group**

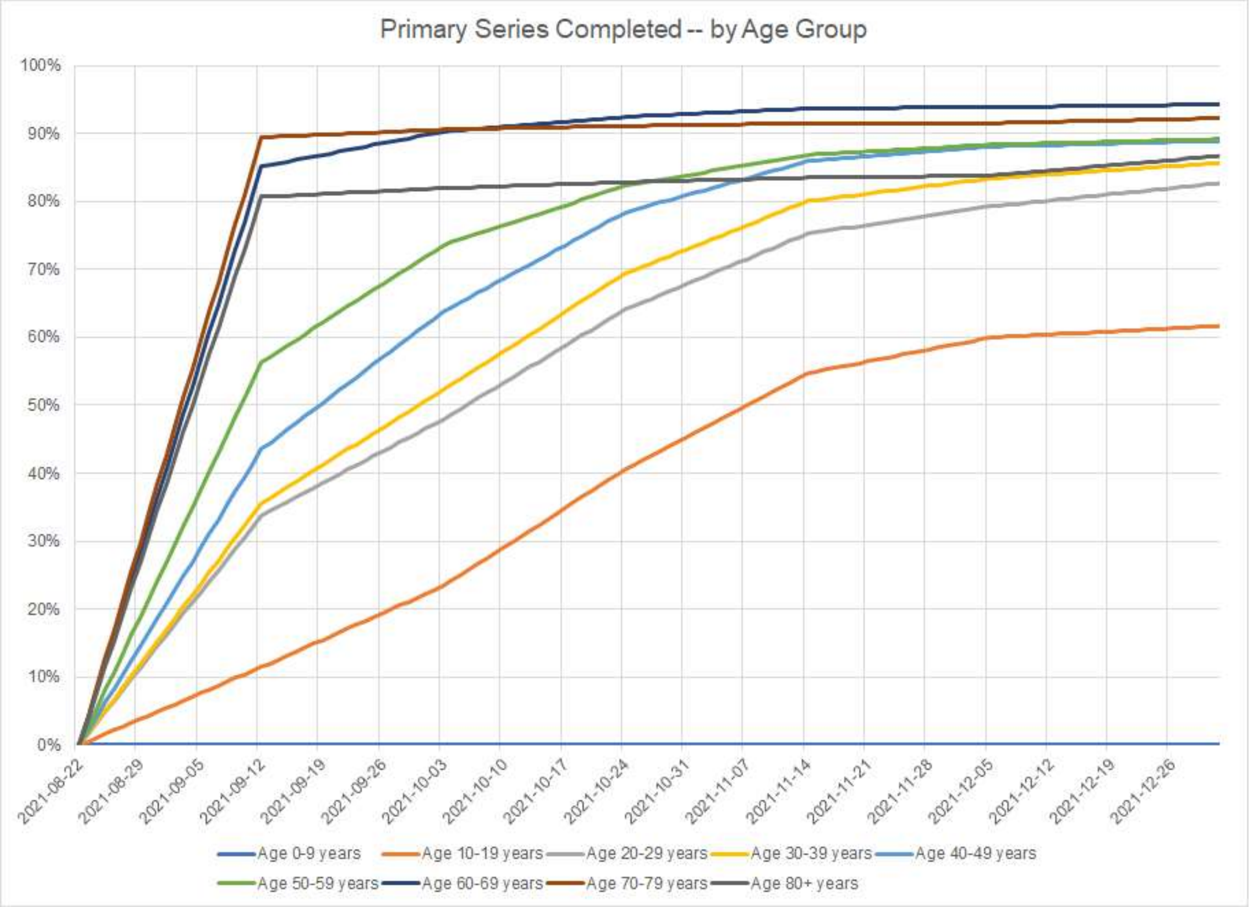

**Figure 4. Percent of the population who have received a booster, by age group**

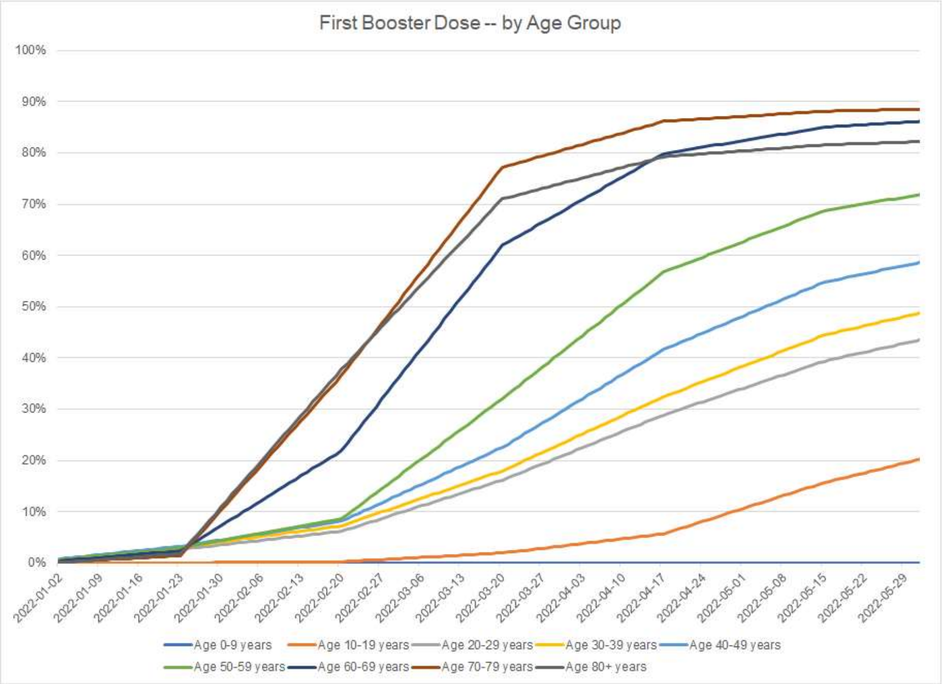

**Figure 5. Percent of the population who received two boosters, by age group**

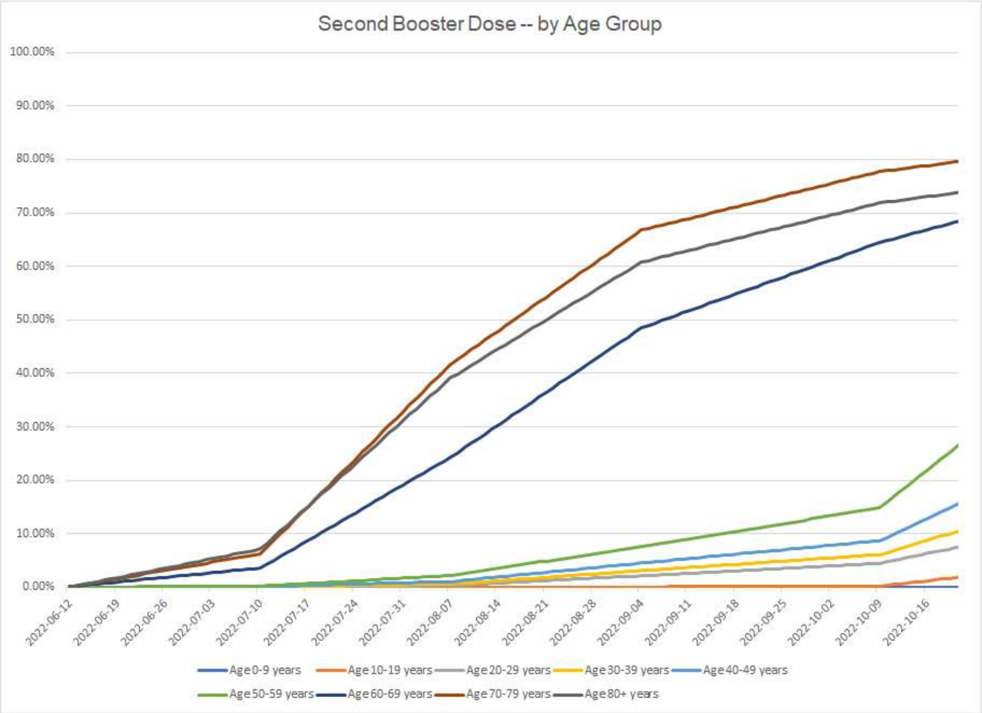

**Figure 6. Percent of the population who received a bivalent booster, by age group**

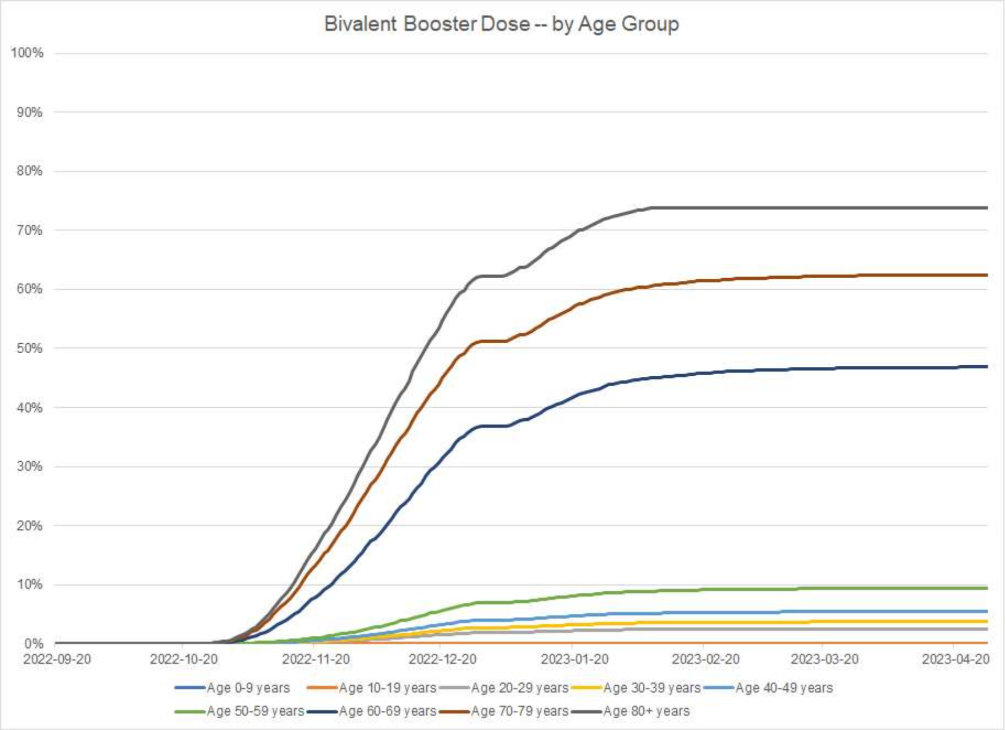

##### 2.3.2. Vaccine Effectiveness

To determine the residual VE (vaccine effectiveness) of the target population, the initial VE and monthly waning for infection and severe disease for vaccines previously received by the population were required. These values differed by the vaccine, vaccination type (primary series or booster), and the circulating variant at the time of administration. Values used are provided below.

<u>Delta</u>

Primary series VE against the Delta variant for infection were obtained from the VERSUS study, which represented the VE of the vaccines used in Japan.[11] As the VERSUS study did not have data on primary series VE against hospitalization, or VE waning for infection and hospitalization for the Delta variant, these were obtained from the Institute for Health Metrics and Evaluation (IHME) COVID-19 December 22, 2021 model update, which provided VE against hospitalization, and waning for infection and hospitalization by individual vaccines.[12] To obtain average values to enter into the model, individual vaccine VEs were weighed by Japanese market shares for

Moderna, Pfizer, and Astra Zeneca so that values would be representative of the mix of vaccines used in Japan.[9]

Boosters were not available when the Delta variant was dominant, and therefore, values were not required

BA.1/BA.2

Primary series and monovalent booster VEs against the BA.1/BA.2 variants for infection and hospitalization were obtained from the 5th and 8th VERSUS study reports,[13,14] respectively. Likewise, monovalent booster VEs against infection and hospitalization were obtained from the 7th and 8th VERSUS study reports.[14,15]. VE waning was obtained from a meta-analysis on the change in duration of booster protection against omicron for symptomatic and severe disease.[16] These values were assumed to be the same for primary series and booster.

BA.4/BA.5

Monovalent booster VE against BA.4/BA.5 for infection was obtained from the VERSUS study 7th report. As no data were available for the protection against severe disease, the proportion decrease observed in VE against infection for BA.1/BA.2 compared to BA.4/BA.5 was applied to the VE against BA.1/BA.2 for severe disease to approximate a value for BA.4/BA.5. Bivalent booster VE was obtained from the VERSUS study data presented at the Japanese Association for Infectious Diseases (JAID) and Scientific Lecture conference.[17] Although the 9th VERSUS study report contains updated data, the time period included in the 9th report extended into March 2023, when the dominant variant had already changed. Therefore, a more accurate estimate of the VE against BA.4/BA.5 presented during the JAID conference, which contained at least a month less of data with the new variant. Waning for both monovalent and bivalent boosters were assumed to be the same as against BA.1/BA.2.[16]

These values were entered into the SEIR model (Table 3).

**Table 3. Starting vaccine effectiveness and monthly waning values for previously administered vaccinations.**

| | Infection | | Severe | |
|---|---|---|---|---|
| | VE | Waning | VE | Waning |
| **DELTA** | | | | |
| Primary series | 88.7% | 3.9% | 95.3% | 1.3% |
| **BA.1/BA.2** | | | | |
| Primary series | 35.6% | 4.8% | 58.2% | 1.4% |
| Monovalent booster | 68.7% | 4.8% | 72.8% | 1.4% |
| **BA.4/BA.5** | | | | |
| Monovalent booster | 50.9% | 4.8% | 53.9% | 1.4% |
| Bivalent booster | 54.7% | 4.8% | 84.9% | 1.4% |

VE: Vaccine effectiveness

On the day that the Omicron BA.1 was emergent (November 11, 2021) and Omicron BA.4/BA.5 was emergent (June 6, 2022), it was assumed that protection from both vaccine-mediated and natural immunity dropped immediately. The decrease was calculated as the ratio of the vaccine effectiveness during each period.

#### 2.4. Other Model Inputs

The average length of time spent by infected individuals exposed to the virus before they become infectious (e.g. latent period) was assumed to be 3 days.[18] The length of time spent by an individual in an infectious state (e.g. infectious period) was assumed to be 7 days.[18,19]

In the base case, the rate of natural immunity waning is set to be equal to the rate of waning of primary series VE against infection. Therefore, the monthly rate of waning is 3.9% for the pre-Omicron period and 4.8% for the Omicron and Omicron BA.4/5 periods. When vaccine-mediated immunity decreased on the days that the circulating variant changed (start of the Omicron period; start of the Omicron BA.4/5 period), a similar drop in the number of people in the Recovered compartment was also implemented.

#### 2.5. Transmissibility: February 2020 to August 2023

During the first 30 days of the burn-in period, an initial 11,493 infections was used to seed or start the pandemic. This number is equal to the number of infections estimated by the Institute for Health Metrics and Evaluation (IHME) over the first 30 days.[20] Model calibration was then conducted to estimate the transmissibility parameter that reflects cases of COVID-19 experienced in Japan from February 2020 through August 2023. The analytical choices for the calibration process were made considering the recommendations of Vanni and colleagues.[21] The transmissibility parameter was allowed to vary on a daily basis. For the calibration process, transmissibility parameters were manually varied. The calibration targets and the goodness of fit measures used, are described along with the results of the calibration below.

##### 2.5.1. Calibration Targets

As the collection of COVID-19 related infections has changed over time, two different data sources for the outcomes of interest, or the target, were used for the calibration process.

For the first part of the pandemic, the target was the total number of infections in the population. This includes all asymptomatic and symptomatic infections, whether or not they have been reported. While data on the number of infections are collected by public health authorities, these figures reflect the number of reported infections and not the true number of infections. There are multiple factors that lead to under-testing and under-reporting of infections including patient and physician behavior, as well as technical limitations to testing. Therefore, output from the modelling done by the IHME, which has been corrected for these biases and includes all infections (symptomatic and asymptomatic), was used as the calibration target. The IHME model was chosen because it was one of the models considered by the United States Centers for Disease Control over time in their ensemble model forecasts.[22] Prior to the pandemic, IHME had developed methods of collecting data globally, which they were able to do in real time during the pandemic in order to create projections for all countries across the globe. IHME has produced multiple publications on their COVID-19 model and the structural and input modifications over time.[23] Finally, the detailed outputs from their model are publicly available. The calibration target was the daily incidence of reported COVID-19 cases (symptomatic and asymptomatic), as reported by the IHME during the period of February 4, 2020 through May 30, 2022.[3] While IHME reported outputs after May 30, 2022, the overall reporting of infections became less reliable overall. Therefore, the endpoint used for this calibration changed after this date.

For the time period June 1, 2022 through May 8, 2023, the calibration target was estimated based on COVID-19 infection data from the Tokyo region.[24] These were inflated to account for the unreported asymptomatic infections and to represent the entire Japanese population. The number of infections was multiplied by 2 to account for reporting bias where those with minor symptomatic infections did not seek medical care or testing for their infections.

The transmissibility parameters were changed manually to estimate the target number of infections. The overall number of infections estimated by IHME, based on the Tokyo data, and the calibration targets adopted are presented in Figure 7.

**Figure 7. Calibration target: Number of COVID-19 infections.**

The number of total infections estimated by IHME and using data collected in Tokyo are shown on the graph with the blue and orange lines respectively. The yellow line represents the calibration target adopted for the base case scenario.

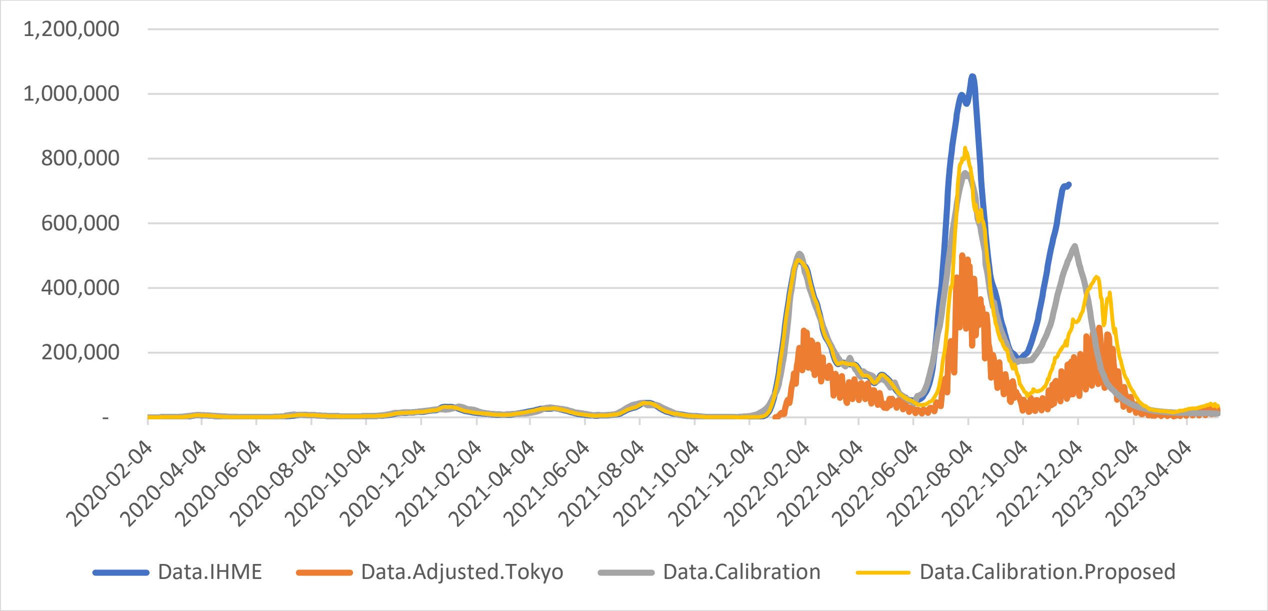

##### 2.5.2. Goodness of Fit Measure

An initial model calibration was performed in a qualitative way by visually comparing the daily incidence of COVID cases predicted by the model and the values obtained from the IHME data or the adjusted Tokyo infection data for the calibration time period.

##### 2.5.3. Results of the Calibration

The final transmissibility parameters for the calibration period for the base case scenario are summarized in Table 4. For the dates shown, the transmissibility parameter was manually varied and linear interpolation was used to estimate values for the days between these dates. The comparison plot of the daily number of incident cases of COVID-19 predicted from the calibrated model and the corresponding calibration targets is presented in Figure 8.

Table 4. Final transmissibility parameter inputs for the SEIR model

| Date | Days from start of analysis | Transmissibility parameter estimate |
|---|---|---|
| January 31, 2020 | 1 | 0.10 |
| February 4, 2020 | 5 | 0.16 |
| February 9, 2020 | 10 | 0.18 |
| February 14, 2020 | 15 | 0.22 |
| February 29, 2020 | 30 | 0.20 |
| March 15, 2020 | 45 | 0.26 |
| March 23, 2020 | 53 | 0.22 |
| March 31, 2020 | 61 | 0.16 |
| April 15, 2020 | 76 | 0.12 |
| April 30, 2020 | 91 | 0.11 |
| May 31, 2020 | 122 | 0.10 |
| June 8, 2020 | 130 | 0.12 |
| June 28, 2020 | 150 | 0.18 |
| June 30, 2020 | 152 | 0.20 |
| July 3, 2020 | 155 | 0.24 |
| July 4, 2020 | 156 | 0.24 |
| July 5, 2020 | 157 | 0.24 |
| July 6, 2020 | 158 | 0.38 |
| July 8, 2020 | 160 | 0.29 |
| July 10, 2020 | 162 | 0.29 |
| July 18, 2020 | 170 | 0.16 |
| July 28, 2020 | 180 | 0.12 |
| July 31, 2020 | 183 | 0.12 |
| August 5, 2020 | 188 | 0.12 |
| August 9, 2020 | 192 | 0.12 |
| August 17, 2020 | 200 | 0.10 |
| August 27, 2020 | 210 | 0.10 |
| August 31, 2020 | 214 | 0.10 |
| September 30, 2020 | 244 | 0.13 |
| October 6, 2020 | 250 | 0.15 |
| October 16, 2020 | 260 | 0.15 |
| October 31, 2020 | 275 | 0.20 |
| November 5, 2020 | 280 | 0.20 |
| November 10, 2020 | 285 | 0.15 |
| November 15, 2020 | 290 | 0.15 |
| November 25, 2020 | 300 | 0.12 |
| November 30, 2020 | 305 | 0.15 |
| December 31, 2020 | 336 | 0.15 |
| January 14, 2021 | 350 | 0.20 |

| Date | Days from start of analysis | Transmissibility parameter estimate |
|---|---|---|
| January 24, 2021 | 360 | 0.13 |
| January 31, 2021 | 367 | 0.13 |
| February 8, 2021 | 375 | 0.10 |
| February 13, 2021 | 380 | 0.10 |
| February 18, 2021 | 385 | 0.10 |
| February 28, 2021 | 395 | 0.12 |
| March 5, 2021 | 400 | 0.10 |
| March 10, 2021 | 405 | 0.15 |
| March 15, 2021 | 410 | 0.15 |
| March 20, 2021 | 415 | 0.15 |
| March 25, 2021 | 420 | 0.15 |
| March 31, 2021 | 426 | 0.15 |
| April 9, 2021 | 435 | 0.18 |
| April 19, 2021 | 445 | 0.21 |
| April 30, 2021 | 456 | 0.17 |
| May 24, 2021 | 480 | 0.10 |
| June 3, 2021 | 490 | 0.10 |
| June 13, 2021 | 500 | 0.08 |
| June 18, 2021 | 505 | 0.12 |
| June 23, 2021 | 510 | 0.12 |
| June 30, 2021 | 517 | 0.14 |
| July 3, 2021 | 520 | 0.15 |
| July 8, 2021 | 525 | 0.18 |
| July 13, 2021 | 530 | 0.26 |
| July 18, 2021 | 535 | 0.26 |
| July 23, 2021 | 540 | 0.24 |
| July 31, 2021 | 548 | 0.19 |
| August 7, 2021 | 555 | 0.18 |
| August 12, 2021 | 560 | 0.15 |
| August 22, 2021 | 570 | 0.15 |
| August 31, 2021 | 579 | 0.13 |
| September 6, 2021 | 585 | 0.13 |
| September 11, 2021 | 590 | 0.12 |
| September 16, 2021 | 595 | 0.10 |
| September 21, 2021 | 600 | 0.10 |
| October 1, 2021 | 610 | 0.11 |
| October 11, 2021 | 620 | 0.12 |
| October 21, 2021 | 630 | 0.16 |
| October 26, 2021 | 635 | 0.16 |
| October 31, 2021 | 640 | 0.16 |
| November 22, 2021 | 662 | 0.35 |
| November 30, 2021 | 670 | 0.30 |
| December 15, 2021 | 685 | 0.37 |
| December 22, 2021 | 692 | 0.37 |
| December 30, 2021 | 700 | 0.39 |
| January 2, 2022 | 703 | 0.35 |
| January 5, 2022 | 706 | 0.34 |
| January 9, 2022 | 710 | 0.39 |
| January 14, 2022 | 715 | 0.39 |

| Date | Days from start of analysis | Transmissibility parameter estimate |
|---|---|---|
| January 16, 2022 | 717 | 0.39 |
| January 18, 2022 | 719 | 0.38 |
| January 19, 2022 | 720 | 0.36 |
| January 24, 2022 | 725 | 0.30 |
| January 29, 2022 | 730 | 0.25 |
| February 3, 2022 | 735 | 0.20 |
| February 8, 2022 | 740 | 0.18 |
| February 14, 2022 | 746 | 0.17 |
| February 28, 2022 | 760 | 0.16 |
| March 15, 2022 | 775 | 0.17 |
| March 20, 2022 | 780 | 0.18 |
| March 25, 2022 | 785 | 0.23 |
| March 31, 2022 | 791 | 0.19 |
| April 2, 2022 | 793 | 0.19 |
| April 4, 2022 | 795 | 0.18 |
| April 6, 2022 | 797 | 0.18 |
| April 8, 2022 | 799 | 0.20 |
| April 9, 2022 | 800 | 0.22 |
| April 19, 2022 | 810 | 0.22 |
| April 24, 2022 | 815 | 0.20 |
| April 29, 2022 | 820 | 0.24 |
| May 1, 2022 | 822 | 0.24 |
| May 4, 2022 | 825 | 0.24 |
| May 9, 2022 | 830 | 0.20 |
| May 14, 2022 | 835 | 0.25 |
| May 19, 2022 | 840 | 0.17 |
| May 24, 2022 | 845 | 0.17 |
| May 29, 2022 | 850 | 0.17 |
| May 31, 2022 | 852 | 0.17 |
| June 2, 2022 | 854 | 0.17 |
| June 5, 2022 | 857 | 0.17 |
| June 8, 2022 | 860 | 0.18 |
| June 15, 2022 | 867 | 0.22 |
| June 23, 2022 | 875 | 0.26 |
| June 30, 2022 | 882 | 0.32 |
| July 3, 2022 | 885 | 0.40 |
| July 15, 2022 | 897 | 0.50 |
| July 23, 2022 | 905 | 0.40 |
| July 31, 2022 | 913 | 0.35 |
| August 2, 2022 | 915 | 0.32 |
| August 7, 2022 | 920 | 0.30 |
| August 12, 2022 | 925 | 0.29 |
| August 14, 2022 | 927 | 0.28 |
| August 17, 2022 | 930 | 0.29 |
| August 22, 2022 | 935 | 0.30 |
| August 27, 2022 | 940 | 0.26 |
| August 31, 2022 | 944 | 0.24 |
| September 6, 2022 | 950 | 0.22 |
| September 11, 2022 | 955 | 0.20 |
| September 16, 2022 | 960 | 0.20 |
| September 26, 2022 | 970 | 0.20 |

| Date | Days from start of analysis | Transmissibility parameter estimate |
|---|---|---|
| October 1, 2022 | 975 | 0.22 |
| October 6, 2022 | 980 | 0.25 |
| October 16, 2022 | 990 | 0.32 |
| October 26, 2022 | 1000 | 0.35 |
| November 5, 2022 | 1010 | 0.48 |
| November 10, 2022 | 1015 | 0.48 |
| November 15, 2022 | 1020 | 0.44 |
| November 23, 2022 | 1028 | 0.42 |
| December 10, 2022 | 1045 | 0.42 |
| December 18, 2022 | 1053 | 0.44 |
| December 20, 2022 | 1055 | 0.45 |
| December 26, 2022 | 1061 | 0.42 |
| January 1, 2023 | 1067 | 0.32 |
| January 2, 2023 | 1068 | 0.40 |
| January 9, 2023 | 1075 | 0.49 |
| January 14, 2023 | 1080 | 0.32 |
| January 19, 2023 | 1085 | 0.30 |
| January 24, 2023 | 1090 | 0.28 |
| January 29, 2023 | 1095 | 0.26 |
| February 1, 2023 | 1098 | 0.22 |
| February 28, 2023 | 1125 | 0.21 |
| March 5, 2023 | 1130 | 0.20 |
| March 31, 2023 | 1156 | 0.40 |
| April 30, 2023 | 1186 | 0.30 |
| May 31, 2023 | 1217 | 0.32 |

**Figure 8 Comparison of daily incidence of COVID cases (calibrated model estimates vs. IHME estimate)**

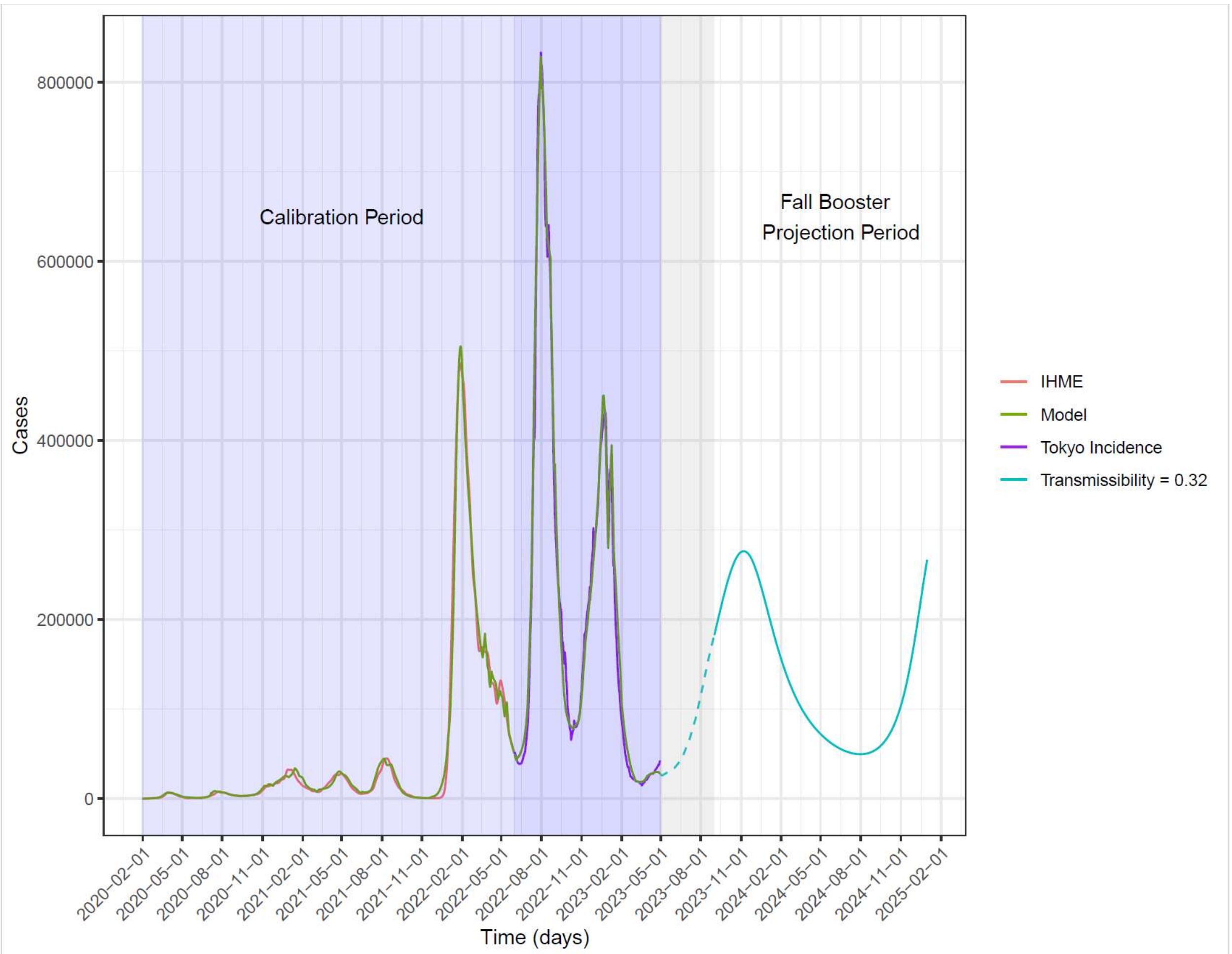

IHME: Institute for Health Metrics and Evaluation

#### 3. SEIR MODEL INPUTS: ANALYTIC TIME PERIOD (SEPTEMBER 2023 TO AUGUST 2024)

The inputs that were changed for the analytic time period are described below. Inputs not described were the same as during the burn-in period.

#### 3.1. Vaccine Coverage

The estimated uptake of a Fall 2023 COVID-19 vaccine was estimated based on observed rates for the first COVID-19 booster in Japan and is displayed by age groups over time in Figure 9.

**Figure 9 Estimated vaccine coverage for a Fall 2023 COVID-19 vaccine**

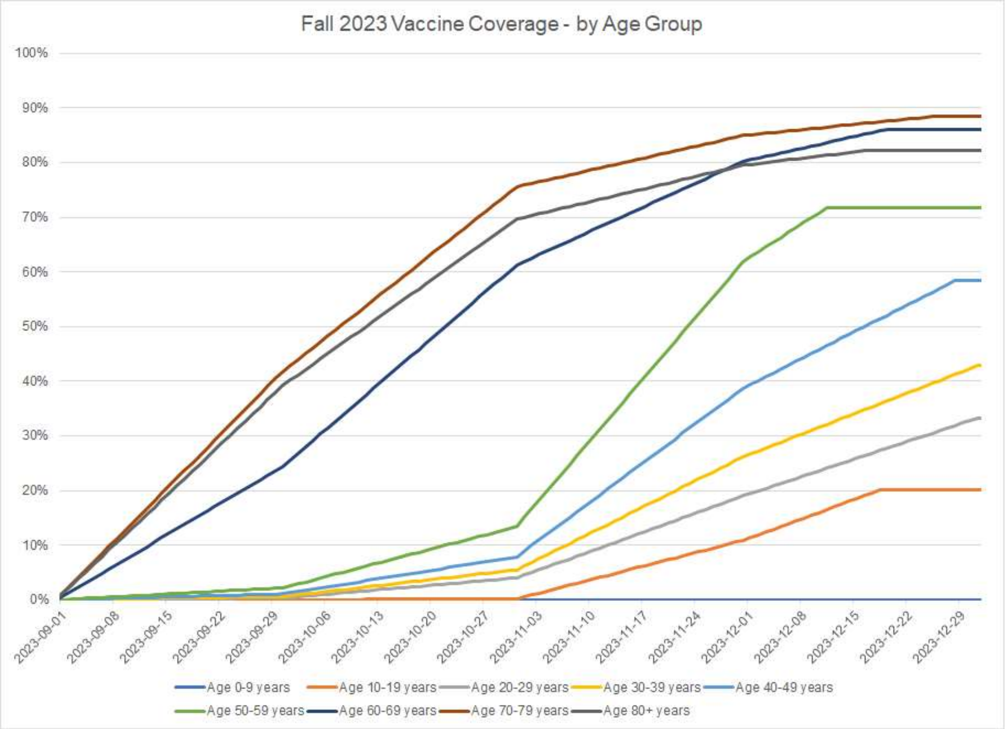

#### 3.2. Vaccine Effectiveness

The estimated effectiveness of the Fall 2023 vaccine was described in the main manuscript. A summary of the inputs used for base case and sensitivity analyses is displayed in Table 5.[14-17,25]

Table 5. Vaccine effectiveness inputs for the Fall 2023 COVID-19 vaccine: base case and sensitivity analyses.

| Analysis | Infection | | Severe Disease | |
|---|---|---|---|---|
| | VE | Waning | VE | Waning |
| Base case | 55% | 4.8% | 85% | 1.4% |
| Range for Sensitivity analysis | 40%-70% | 3.1%-6.8% | 75%-95% | 0.6%-2.4% |

##### 3.3. Transmissibility: April 1, 2023 onwards

The values for the transmissibility parameter after September 1, 2023, were held constant at the final value in the calibration period (0.32) for the projections, assuming it represents the transmissibility of the variability of the currently circulating variant.

#### 4. SEIR MODEL SCENARIO ANALYSES

Several scenario analyses were developed as described in the manuscript to show the impact of different incidence projections on predicted COVID-19 burden. Three new calibrations were conducted and the new targets and the match of the model predicted outcomes is shown along with base case in Figure 10 Comparison of daily incidence of COVID-19 cases (calibrated model estimates versus targets). Panel A represents base case, Panel B to Panel D represent different calibration scenarios. For one of the scenarios, the mobility scalar was changed as shown in Figure 11, where it is compared to base case. The transmissibility parameters used for each of the incidence scenarios is displayed in Table 8. The projected incidence for each of the scenarios is shown in Figure 12.

Table 6. Transmissibility parameter used for the analysis period for the different incidence scenario analyses.

| Scenario | Transmissibility Parameter |
|---|---|
| Base case | 0.32 |
| Immune Escape April 2024 | 0.32 |
| Immune Escape June 2024 | 0.32 |
| Adjusted Tokyo Data (2.5x) | 0.45 |
| Adjusted Tokyo Data (1.5x) | 0.25 |
| Seasonality: phi=0.2 | 0.32 |
| Adjusted Tokyo Data (2.0x), Revised Waning* | 0.30 |
| Lower transmissibility | 0.25 |
| Higher transmissibility | 0.45 |

**Figure 10 Comparison of daily incidence of COVID-19 cases (calibrated model estimates versus targets). Panel A represents base case, Panel B to Panel D represent different calibration scenarios.**

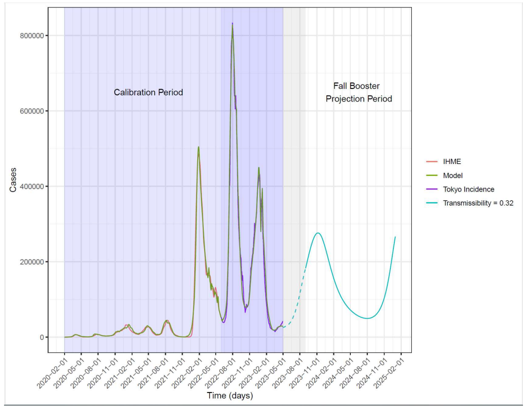

Panel A: Base case (Infection targets multiplied by 2)

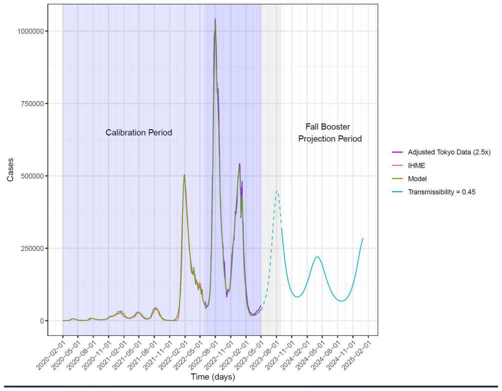

Panel C: Base case (Infection targets multiplied by 2.5)

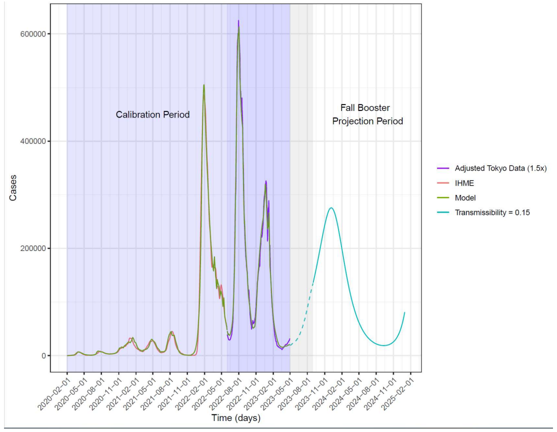

Panel B: Scenario: Infection targets multiplied by 1.5

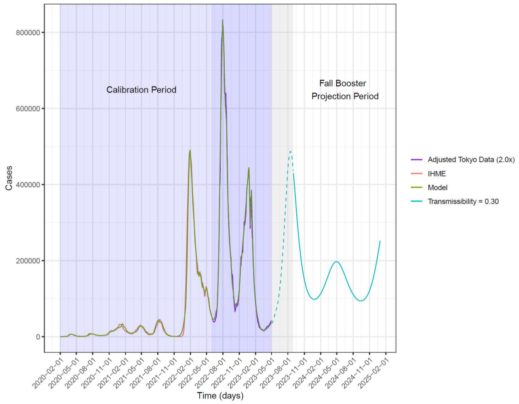

Panel D: Scenario: Infection targets multiplied by 2; Vaccine waning: 6.58%

IHME: Institute for Health Metrics and Evaluation

**Figure 11 Mobility scaling factor applied to the contact matrix to change contact patterns over time: base case and scenario analysis.**

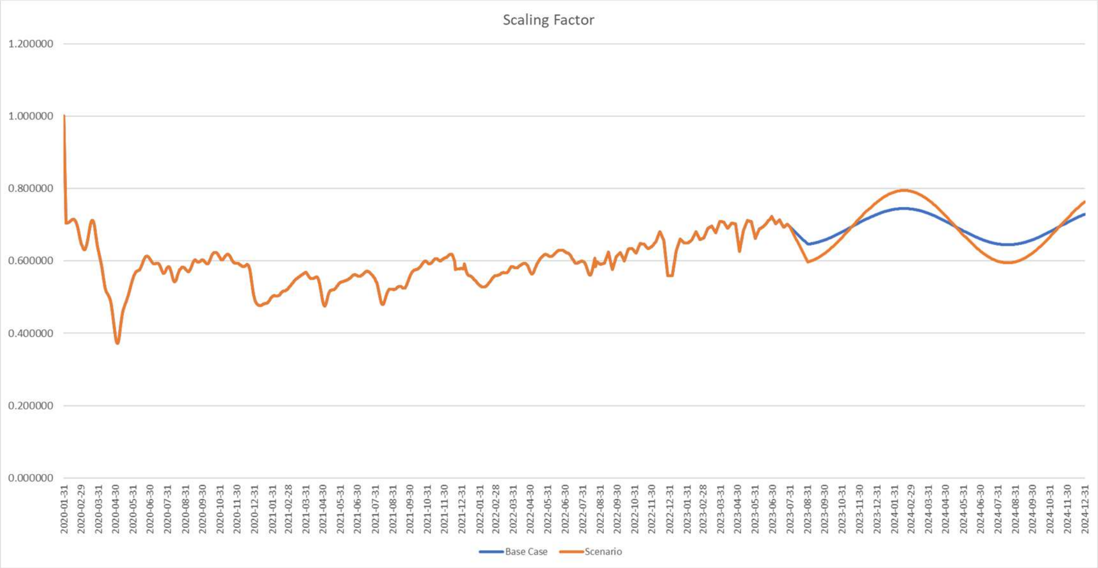

**Figure 12 Impact of varying SEIR model inputs on COVID-19 incidence projections (symptomatic infections) without a Fall 2023 COVID-19 Vaccine**

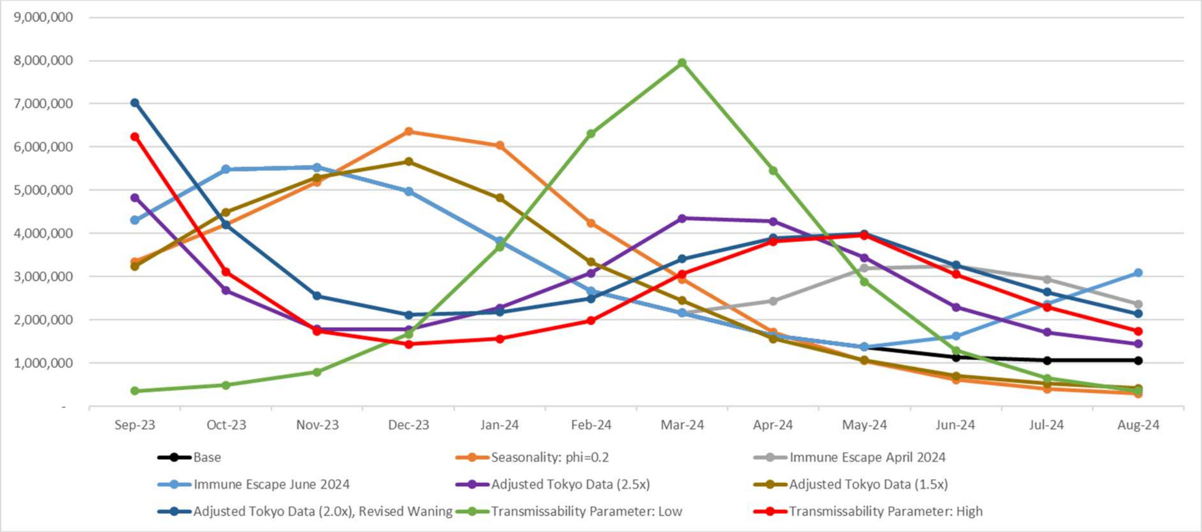

#### 5. CONSEQUENCES OF INFECTION: DETAILS OF INPUTS

##### 5.1. Hospitalization rates

The hospitalization rates amongst those with symptomatic COVID-19 infections in an unvaccinated population were estimated based on observed hospitalization data, assumed vaccination rates, and assumed underlying protection against hospitalizations in the vaccinated population. The observed COVID-19 hospitalization rates, by age group, (column 2 of Table 7) were derived from an analysis of the NIID data. They are similar to hospitalization rates from two difference insurance databases: JAST Employee Insurance data and the DeSC Insurance data. The hospitalization rates are presented as the number of COVID-19 hospitalizations divided by the number of COVID-19 cases. By definition, the denominator is cases that have come to medical attention and received some sort of medical care, if only consultation with a medical professional. The proportion of each age group that was assumed to be vaccinated was assumed to be the same as the uptake of the primary series in the general population. These data were obtained from vaccine uptake data from the Ministry of Health, Labour and Welfare of Japan website and the Official Website of the Prime Minister of Japan and His Cabinet during the period April 1, 2022 through March 31, 2023 (column 4 of Table 7). [9,10] The proportion of each age group that were assumed to be unvaccinated was calculated as 100% minus the proportion vaccinated (column 3 of Table 7). This is likely an under-estimate as it is expected that a higher proportion in the hospitalized population are unvaccinated. Finally, we assumed that the residual VE in the vaccinated population against hospitalization was 40%. This residual VE would represent the average VE in the population with a mixed vaccination history (i.e., primary doses; booster doses 1 – 3), across the one year period. The hospitalization rates in an unvaccinated population with medically attended, symptomatic infections were estimated using the equations below. We assume the observed hospitalization rates in those with medically attended, symptomatic infections (column 2 of Table 7) are a weighted average of the rates in an unvaccinated population and vaccinated population weighted by the proportion unvaccinated and vaccinated in the population. For each age group, the following equation was used to estimate the hospitalization rate in an unvaccinated population.

As the model requires hospitalization rates amongst all symptomatic cases and not just those that are medically attended, the calculated rates were adjusted. For our base case calibration, we assumed that 50% of cases were medically attended. Therefore, the rates were multiplied by 50% and the final inputs are shown in the last column of Table 8.

$$Hospitalization\ Rate_{Observed} = \left[(\%\ Unvaccinated) \times Hospitalization\ Rate_{Unvaccinated}\right] + \left[(\%\ Vaccinated) \times Hospitalization\ Rate_{Vaccinated}\right]$$

$$Hospitalization\ Rate_{Observed} = \left[(\%\ Unvaccinated) \times Hospitalization\ Rate_{Unvaccinated}\right] + \left[(100\% - \ \%\ Unvaccinated) \times (Hospitalization\ Rate_{Unvaccinated} \times (1 - VE)\right]$$

By substituting in $\%\ Vaccinated = 100\% - \ \%\ Unvaccinated$ and

$$Hospitalization\ Rate_{Vaccinated} = \ Hospitalization\ Rate_{Unvaccinated} \times (1 - VE)$$

Rearranging yields:

$$Hospitalization\ Rate_{Unvaccinated} = \frac{Hospitalization\ Rate_{Observed}}{(\%\ Vaccinated) + [(100\% - \ \%\ Unvaccinated) \times (1 - VE)]}$$

$$Hospitalization\ Rate_{Unvaccinated,\ \ Symptomatic\ Disease} = (\%\ Cases\ Medically\ attended) \times Hospitalization\ Rate_{Unvaccinated}$$

**Table 7. Hospitalization Rates.**

| Age Group | Hospitalization Rate (Observed)* | Proportion Assumed Unvaccinated** | Proportion Vaccinated*** | Residual VE Hospitalization in Vaccinated Population | Hospitalization Rate (Unvaccinated Population)* | Hospitalization Rate (Unvaccinated; Symptomatic Disease)**** |
|---|---|---|---|---|---|---|
| 0-11 years | 0.92% | 100.0% | 0.0% | 40.0% | 0.92% | 0.46% |
| 12-17 years | 0.26% | 38.3% | 61.7% | 40.0% | 0.35% | 0.18% |
| 18-29 years | 0.40% | 17.4% | 82.6% | 40.0% | 0.60% | 0.30% |
| 30-39 years | 0.62% | 14.4% | 85.6% | 40.0% | 0.94% | 0.47% |
| 40-49 years | 0.65% | 11.1% | 88.9% | 40.0% | 1.01% | 0.51% |
| 50-59 years | 1.34% | 10.8% | 89.2% | 40.0% | 2.08% | 1.04% |
| 60-64 years | 3.65% | 5.8% | 94.3% | 40.0% | 5.86% | 2.93% |
| 65-74 years | 10.81% | 6.8% | 93.2% | 40.0% | 17.25% | 8.62% |
| 75+ years | 17.98% | 13.4% | 86.7% | 40.0% | 27.51% | 13.76% |

* Denominator: All symptomatic cases that have come to medical attention.
** Assumed based on rates of uptake of primary series in the general population.
***Proportion vaccinated = 100% - proportion unvaccinated.
****Assumed equal to Hospitalization Rate (Unvaccinated Population multiplied by 50% of cases medically attended
VE: vaccine effectiveness

##### 5.2. Mortality rates

The mortality rates were derived from an analysis of two difference insurance databases: JAST Employee Insurance data (for those >60 years) and the DeSC Citizen's Insurance & Superelderly (for those 60 years and older). The case definition was death within 3 months of hospitalization with a COVID-19 diagnoses. One of the limitations is that those who were hospitalized with COVID-19 for more than 3 months and then died would not be counted in this analysis, leading to some underestimation. On the other hand, those who were hospitalized with COVID-19 who died from other causes would be counted as a COVID-19 in-hospital deaths, leading to some overestimation.

#### 6. ADDITIONAL RESULTS TABLES AND FIGURES

**Table 8 Scenario analyses: projected number of symptomatic infections, hospitalizations, and deaths with and without a COVID-19 vaccine updated for Fall 2023.**

| | Symptomatic infections | | | Hospitalizations | | | Deaths | | |
|---|---|---|---|---|---|---|---|---|---|
| **Scenario** | **No Vaccine** | **With Vaccine** | **Prevented** | **No Vaccine** | **With Vaccine** | **Prevented** | **No Vaccine** | **With Vaccine** | **Prevented** |
| **Base case** | 35,240,923 | 27,915,148 | 7,325,775 | 689,973 | 415,239 | -274,734 | 61,738 | 35,881 | -25,857 |
| **Target Population** | | | | | | | | | |
| 65 years + only | 35,240,923 | 32,337,129 | 2,903,794 | 689,973 | 509,560 | 180,413 | 61,738 | 42,266 | 19,472 |
| **Hospitalization rates** | | | | | | | | | |
| Higher hospitalization rates | 35,240,923 | 27,915,148 | 7,325,775 | 799,165 | 480,102 | 319,063 | 71,553 | 41,579 | 29,975 |
| Lower hospitalization rates | 35,240,923 | 27,915,148 | 7,325,775 | 607,486 | 366,200 | 241,286 | 54,297 | 31,561 | 22,736 |
| **Initial Vaccine Effectiveness (VE)** | | | | | | | | | |
| Low hospital VE | 35,240,923 | 27,915,148 | 7,325,775 | 689,973 | 493,685 | 196,287 | 61,738 | 43,613 | 18,125 |
| High hospital VE | 35,240,923 | 27,915,148 | 7,325,775 | 689,973 | 336,793 | 353,180 | 61,738 | 28,149 | 33,589 |
| Low infection VE | 35,240,923 | 33,562,227 | 1,678,696 | 689,973 | 481,969 | 208,004 | 61,738 | 42,224 | 19,514 |
| High infection VE | 35,240,923 | 20,329,008 | 14,911,915 | 689,973 | 311,296 | 378,677 | 61,738 | 26,360 | 35,378 |
| **Vaccine Waning** | | | | | | | | | |
| Low waning (infection) | 35,240,923 | 27,915,148 | 7,325,775 | 689,973 | 385,085 | 304,887 | 61,738 | 32,908 | 28,830 |
| High waning (infection) | 35,240,923 | 27,915,148 | 7,325,775 | 689,973 | 455,574 | 234,398 | 61,738 | 39,859 | 21,880 |
| Low waning (hospitalization) | 35,240,923 | 23,026,242 | 12,214,681 | 689,973 | 348,012 | 341,961 | 61,738 | 29,740 | 31,998 |
| High waning (hospitalization) | 35,240,923 | 33,701,803 | 1,539,120 | 689,973 | 491,193 | 198,780 | 61,738 | 42,953 | 18,786 |

| | | | | | | | | | |
|---|---|---|---|---|---|---|---|---|---|
| **Vaccine Coverage** | | | | | | | | | |
| 75% of base case | 35,240,923 | 29,992,292 | 5,248,630 | 689,973 | 484,249 | 205,724 | 61,738 | 42,268 | 19,470 |
| 50% of base case | 35,240,923 | 31,805,501 | 3,435,422 | 689,973 | 552,414 | 137,559 | 61,738 | 48,653 | 13,085 |
| **COVID-19 Incidence** | | | | | | | | | |
| Immune Escape April 2024 | 43,148,725 | 37,965,618 | 5,183,107 | 842,573 | 609,562 | 233,011 | 74,951 | 53,519 | 21,431 |
| Immune Escape June 2024 | 39,068,442 | 34,128,544 | 4,939,899 | 759,749 | 529,928 | 229,821 | 67,622 | 46,113 | 21,508 |
| Adjusted Tokyo Data (2.5x) | 33,985,577 | 32,456,608 | 1,528,969 | 707,888 | 524,135 | 183,753 | 64,335 | 47,176 | 17,159 |
| Adjusted Tokyo Data (1.5x) | 33,606,420 | 22,073,677 | 11,532,743 | 622,411 | 303,637 | 318,774 | 54,581 | 25,375 | 29,206 |
| Seasonality: phi=0.2 | 36,426,098 | 28,744,878 | 7,681,220 | 723,775 | 429,706 | 294,068 | 65,096 | 37,377 | 27,719 |
| Adjusted Tokyo Data (2.0x), Revised Waning* | 39,927,629 | 38,124,689 | 1,802,941 | 888,266 | 610,597 | 277,669 | 78,996 | 53,505 | 25,491 |
| Lower Transmissibility | 31,909,792 | 21,719,169 | 10,190,624 | 605,424 | 287,905 | 317,520 | 52,952 | 24,436 | 28,515 |
| Higher Transmissibility | 34,029,660 | 29,938,732 | 4,090,928 | 710,885 | 484,227 | 226,657 | 64,544 | 43,344 | 21,201 |

* Waning against infection was set to 6.58% during Omicron based on inputs provided by Prof. Igarashi, including historically (during the calibration) and for the Fall 2023 vaccine.

### 7. EQUATIONS ASSOCIATED WITH THE SEIR MODEL

#### 7.1. Differential Equations

The following equations describe the movement of people in the population through the SEIR compartments and the vaccination strata.

**Unvaccinated cohort**

$$S_{t+1,j}^{X} = S_{t,j}^{X} - \lambda_{t,j}^{X} S_{t,j}^{X} - \left(p_{t,j}^{X,S}\right)\mu_{t,j} + \omega_{t,j}^{X} R_{t,j}^{X} - \epsilon_{t,j}$$

$$E_{t+1,j}^{X} = E_{t,j}^{X} + \lambda_{t,j}^{X} S_{t,j}^{X} - \frac{1}{\tau_E} E_{t,j}^{X}$$

$$I_{t+1,j}^{X} = I_{t,j}^{X} + \frac{1}{\tau_E} E_{t,j}^{X} - \frac{1}{\tau_I} I_{t,j}^{X} + \epsilon_{t,j}$$

$$R_{t+1,j}^{X} = R_{t,j}^{X} - \left(p_{t,j}^{X,R}\right)\mu_{t,j} + \frac{1}{\tau_I} I_{t,j}^{X} - \omega_{t,j}^{X} R_{t,j}^{X}$$

**Vaccinated cohort**

$$S_{t+1,j}^{V} = S_{t,j}^{V} - \lambda_{t,j}^{V} S_{t,j}^{V} + \left(p_{t,j}^{X,S}\right)\mu_{t,j} - \left(p_{t,j}^{V,S}\right)\nu_{t,j} + \omega_{t,j}^{V} R_{t,j}^{V}$$

$$E_{t+1,j}^{V} = E_{t,j}^{V} + \lambda_{t,j}^{V} S_{t,j}^{V} - \frac{1}{\tau_E} E_{t,j}^{V}$$

$$I_{t+1,j}^{V} = I_{t,j}^{V} + \frac{1}{\tau_E} E_{t,j}^{V} - \frac{1}{\tau_I} I_{t,j}^{V}$$

$$R_{t+1,j}^{V} = R_{t,j}^{V} + \left(p_{t,j}^{X,R}\right)\mu_{t,j} - \left(p_{t,j}^{V,R}\right)\nu_{t,j} + \frac{1}{\tau_I} I_{t,j}^{V} - \omega_{t,j}^{V} R_{t,j}^{V}$$

**First Booster cohort**

$$S_{t+1,j}^{B} = S_{t,j}^{B} - \lambda_{t,j}^{B} S_{t,j}^{B} + \left(p_{t,j}^{V,S}\right)\nu_{t,j} - \left(p_{t,j}^{B,S}\right)\nu_{t,j}^{2} - \left(p_{t,j}^{B,S}\right)\left(p_{t,j}^{B,B4}\right)\nu_{t,j}^{4} + \omega_{t,j}^{B} R_{t,j}^{B}$$

$$E_{t+1,j}^{B} = E_{t,j}^{B} + \lambda_{t,j}^{B} S_{t,j}^{B} - \frac{1}{\tau_E} E_{t,j}^{B}$$

$$I_{t+1,j}^{B} = I_{t,j}^{B} + \frac{1}{\tau_E} E_{t,j}^{B} - \frac{1}{\tau_I} I_{t,j}^{B}$$

| |
|---|
| $$R^{B}_{t+1,j} = R^{B}_{t,j} + \left(p^{V,R}_{t,j}\right)v_{t,j} - \left(p^{B,R}_{t,j}\right)v^{2}_{t,j} - \left(p^{B,R}_{t,j}\right)\left(p^{B,B4}_{t,j}\right)v^{4}_{t,j} + \frac{1}{\tau_I}I^{B}_{t,j} - \omega^{B}_{t,j}R^{B}_{t,j}$$ |
| **Second Booster cohort** |
| $$S^{B2}_{t+1,j} = S^{B2}_{t,j} - \lambda^{B2}_{t,j}S^{B2}_{t,j} + \left(p^{B,S}_{t,j}\right)v^{2}_{t,j} - \left(p^{B2,S}_{t,j}\right)v^{3}_{t,j} - \left(p^{B2,S}_{t,j}\right)\left(p^{B2,B4}_{t,j}\right)v^{4}_{t,j} + \omega^{B2}_{t,j}R^{B2}_{t,j}$$ $$E^{B2}_{t+1,j} = E^{B2}_{t,j} + \lambda^{B2}_{t,j}S^{B2}_{t,j} - \frac{1}{\tau_E}E^{B2}_{t,j}$$ $$I^{B2}_{t+1,j} = I^{B2}_{t,j} + \frac{1}{\tau_E}E^{B2}_{t,j} - \frac{1}{\tau_I}I^{B2}_{t,j}$$ $$R^{B2}_{t+1,j} = R^{B2}_{t,j} + \left(p^{B,R}_{t,j}\right)v^{2}_{t,j} - \left(p^{B2,R}_{t,j}\right)v^{3}_{t,j} - \left(p^{B2,R}_{t,j}\right)\left(p^{B2,B4}_{t,j}\right)v^{4}_{t,j} + \frac{1}{\tau_I}I^{B2}_{t,j} - \omega^{B2}_{t,j}R^{B2}_{t,j}$$ |
| **Third Booster cohort** |
| $$S^{B3}_{t+1,j} = S^{B3}_{t,j} - \lambda^{B3}_{t,j}S^{B3}_{t,j} + \left(p^{B2,S}_{t,j}\right)v^{3}_{t,j} - \left(p^{B3,S}_{t,j}\right)\left(p^{B3,B4}_{t,j}\right)v^{4}_{t,j} + \omega^{B3}_{t,j}R^{B3}_{t,j}$$ $$E^{B3}_{t+1,j} = E^{B3}_{t,j} + \lambda^{B3}_{t,j}S^{B3}_{t,j} - \frac{1}{\tau_E}E^{B3}_{t,j}$$ $$I^{B3}_{t+1,j} = I^{B3}_{t,j} + \frac{1}{\tau_E}E^{B3}_{t,j} - \frac{1}{\tau_I}I^{B3}_{t,j}$$ $$R^{B3}_{t+1,j} = R^{B3}_{t,j} + \left(p^{B2,R}_{t,j}\right)v^{3}_{t,j} - \left(p^{B3,R}_{t,j}\right)\left(p^{B3,B4}_{t,j}\right)v^{4}_{t,j} + \frac{1}{\tau_I}I^{B3}_{t,j} - \omega^{B3}_{t,j}R^{B3}_{t,j}$$ |
| **Fourth Booster cohort** |
| $$S^{B4}_{t+1,j} = S^{B4}_{t,j} - \lambda^{B4}_{t,j}S^{B4}_{t,j} + \left(\left(p^{B,S}_{t,j}\right)p^{B,B4}_{t,j} + \left(p^{B2,S}_{t,j}\right)p^{B2,B4}_{t,j} + \left(p^{B3,S}_{t,j}\right)p^{B3,B4}_{t,j}\right)v^{4}_{t,j} + \omega^{B4}_{t,j}R^{B4}_{t,j}$$ $$E^{B4}_{t+1,j} = E^{B4}_{t,j} + \lambda^{B4}_{t,j}S^{B4}_{t,j} - \frac{1}{\tau_E}E^{B4}_{t,j}$$ $$I^{B4}_{t+1,j} = I^{B4}_{t,j} + \frac{1}{\tau_E}E^{B4}_{t,j} - \frac{1}{\tau_I}I^{B4}_{t,j}$$ $$R^{B4}_{t+1,j} = R^{B4}_{t,j} + \left(\left(p^{B,R}_{t,j}\right)p^{B,B4}_{t,j} + \left(p^{B2,R}_{t,j}\right)p^{B2,B4}_{t,j} + \left(p^{B3,R}_{t,j}\right)p^{B3,B4}_{t,j}\right)v^{4}_{t,j} + \frac{1}{\tau_I}I^{B4}_{t,j} - \omega^{B4}_{t,j}R^{B4}_{t,j}$$ |
| **For t=1,** |

| |
|---|
| $S_{1,j}^{X}$ = Initial number of susceptible individuals<br><br>$I_{1,j}^{X}$= Initial number of infectious individuals = $\epsilon_{1,j}$<br><br>$E_{1,j}^{X} = R_{1,j}^{X} = 0$<br><br>$S_{1,j}^{V} = E_{1,j}^{V} = I_{1,j}^{V} = R_{1,j}^{V} = 0$<br><br>$S_{1,j}^{B} = E_{1,j}^{B} = I_{1,j}^{B} = R_{1,j}^{B} = 0$<br><br>$S_{1,j}^{B2} = E_{1,j}^{B2} = I_{1,j}^{B2} = R_{1,j}^{B2} = 0$<br><br>$S_{1,j}^{B3} = E_{1,j}^{B3} = I_{1,j}^{B3} = R_{1,j}^{B3} = 0$<br><br>$S_{1,j}^{B4} = E_{1,j}^{B4} = I_{1,j}^{B4} = R_{1,j}^{B4} = 0$ |
| Notes, superscripts:<br><br>*X*, *V*, *B*, *B2*, *B3*, and *B4* represent the unvaccinated, vaccinated, first booster, second booster, third booster (bivalent), and fourth booster (fall 2023 bivalent) cohorts, respectively.<br><br>Notes, subscripts:<br><br>*t* = time (i.e., day of analysis)<br><br>*j* = age group (number 1 to 9) |
| **Definitions** |
| $S_j^X, S_j^V, S_j^B, S_j^{B2}, S_j^{B3}, S_j^{B4}$ represent the proportion of susceptibles in age group *j* in cohort *X*, *V, B, B2, B3*, or *B4*.<br><br>The compartments with superscript *X*, *V*, *B, B1, B2, B3, and B4* represent the unvaccinated, vaccinated, and boosted cohorts, respectively.<br><br>$E_j^Z$ represent exposed, but not yet infectious, individuals in age group *j* in cohort *Z* (*X*, *V*, *B, B2*, *B3*, or *B4*).<br><br>$I_j^Z$ represent infectious individuals in age group *j* in cohort *Z* (*X*, *V*, *B, B2*, *B3*, or *B4*).<br><br>$R_j^Z$ represent immune individuals in age group *j* in cohort *Z* (*X*, *V*, *B, B2*, *B3*, or *B4*). |
| $\lambda_{t,j}^{*}$ is the age-group specific force of infection (see section below) |

$\frac{1}{\tau_E}$ is the rate of loss of latency

$\frac{1}{\tau_I}$ is the rate of loss of infectiousness

$\mu_{t,j}$ is the proportion receiving a (primary series) vaccination on day *t* in age group *j*

$\nu_{t,j}$ is the proportion receiving a booster on day *t* in age group *j*

$\nu_{t,j}^2$ is the proportion receiving a second booster on day *t* in age group *j*

$\nu_{t,j}^3$ is the proportion receiving a third booster on day *t* in age group *j*

$\nu_{t,j}^4$ is the proportion receiving a fourth booster on day *t* in age group *j*

$p_{t,j}^{B,B4}$ is the proportion receiving a fourth booster from cohort *B* on day *t* in age group *j*

$p_{t,j}^{B2,B4}$ is the proportion receiving a fourth booster from cohort *B2* on day *t* in age group *j*

$p_{t,j}^{B3,B4}$ is the proportion receiving a fourth booster from cohort *B3* on day *t* in age group *j*

Note: $p_{t,j}^{B,B4} + p_{t,j}^{B2,B4} + p_{t,j}^{B3,B4} = 1$

$p_{t-1,j}^{Z,S}$ is the proportion of the booster cohort *Z in the S* compartment out of the total proportion of the booster cohort *Z in the S and R* compartments on day *t-1* in age group *j*

$p_{t-1,j}^{Z,R}$ is the proportion of the booster cohort *Z in the R* compartment out of the total proportion of the booster cohort *Z in the S and R* compartments on day *t-1* in age group *j*

Note: $p_{t-1,j}^{Z,S} + p_{t-1,j}^{Z,R} = 1$

$\omega_{t,j}^Z$ is the natural immunity waning rate on day *t* in age group *j* in cohort Z (*X*, *V*, *B*, *B2*, *B3*, or *B4)*

$\epsilon_{t,j}$ is the proportion of external cases on day *t* in age group *j*

#### 7.2. Force of infection (unvaccinated)

$$\lambda_{t,i}^* = (Overall\ Scaling\ Factor_t) \times \left[ \beta_t \sum_{j=1}^{9} \sum_{Z=\{X,V,B\}} c_{ij} I_{t,j}^Z \right]$$

$\lambda^{*}_{t,i}$ is the age-group specific force of infection at time *t* for age group *i*

$Overall\ \ Scaling\ Factor_t$ at time *t* is defined in section 2.2.2.

$\beta_t$ is the transmissibility parameter at time *t*

$c_{ij}$ is the rate at which individuals in age group *i* make contact with those in age group *j*

$I^{Z}_{t,j}$ represents the infectious individuals in cohort *Z* (*X*, *V*, *B, B2*, *B3*, or *B4*).at time *t* for age group *j*.

##### 7.3. Force of infection (vaccinated)

For the vaccinated cohorts, the force of infection calculation is adjusted based on the VE in the cohort:

$$\lambda^{Z}_{t,i} = \left(1 - VE^{X}_{t,i}\right) \times (Overall\ Scaling\ Factor_t) \times \left[\beta_t \sum_{j=1}^{9} \sum_{Z=\{X,V,B\}} c_{ij} I^{Z}_{t,j}\right]$$

$\lambda^{Z}_{t,i}$ is the age-group specific force of infection at time *t* for age group *i* in cohort *Z* (*X*, *V*, *B, B2*, *B3*, or *B4*).

$VE^{Z}_{t,i}$ is the vaccine effectiveness at time *t* for age group *i* in cohort *Z* (*X*, *V*, *B, B2*, *B3*, or *B4*).The vaccine effectiveness at time *t* is defined in the next section.

##### 7.4. Daily vaccine effectiveness calculations

If no one is vaccinated in the cohort and age group, a VE value of zero is assumed (i.e., when day *t* is less than the day of the beginning of the vaccination period). Once people have been vaccinated in the vaccination cohort and age group, the average vaccine effectiveness on day *t* is calculated as:

$$VE^{Z}_{t,i} = \frac{\left[\begin{array}{c}(Number\ newly\ vaccinated\ on\ day\ t \times initial\ VE) + \\ \Big((VE\ Drop_t)\big(Number\ previously\ vaccinated\ on\ day\ t \times (VE^{Z}_{t-1,i} - daily\ waning\ rate)\big)\Big)\end{array}\right]}{[Total\ number\ in\ cohort\ Z\ on\ day\ t]}$$

$VE^{Z}_{t,i}$ is the vaccine effectiveness at time *t* for age group *i* in cohort *Z* (*X*, *V*, *B, B2*, *B3* or *B4*).

If the term $\left(VE_{t-1,i}^{Z} - daily\ waning\ rate\right)$ falls below zero, we assume a value of zero instead.

The term $(VE\ Drop_t)$ represents a drop in the vaccine effectiveness when a new strain with immune escape enters the population (see Section 2.3.2. There are only a few days in which this drop occurs over the course of the time horizon: if a new variant with immune escape emerges, the impact is assumed to happen on the individual days described in Section 5.5.8. Apart from those days, the value of the term $(VE\ Drop_t)$ is assumed to be one, corresponding to no impact on the average VE calculation.

##### 7.5. Calculation of incremental effectiveness against hospitalization

For each vaccination cohort, age group, and day, we define the following vaccine effectiveness variables and relationship between the variables. The superscripts and subscripts for vaccination cohort, age group, and day are removed for clarity.

**Definitions**

$VE_1$ = Vaccine effectiveness against infection

$VE_2$ = 'Total' Vaccine effectiveness against hospitalization

$VE_2^*$= 'Additional' Vaccine effectiveness against hospitalization

We assume $VE_2^* = 0$ if there is no additional benefit against hospitalization

**Define**

$$[1 - VE_2] = [1 - VE_1] \times [1 - VE_2^*]$$

Isolate and solve for $VE_2^*$

$$[1 - VE_2^*] = \frac{[1 - VE_2]}{[1 - VE_1]}$$

$$VE_2^* = 1 - \frac{[1 - VE_2]}{[1 - VE_1]}$$

**Probabilities in an unvaccinated cohort**

Probability of COVID-19 infection in an unvaccinated cohort

$$p(COVID|UnVac)$$

Probability that a COVID-19 infection requires hospitalization in an unvaccinated cohort

$$p(Hosp|COVID, UnVac)$$

Proportion of an unvaccinated cohort with a COVID-19 infection that requires hospitalization

$$p(Hosp, COVID|UnVac) = p(Hosp|COVID, UnVac) \times p(COVID|UnVac)$$

**Probabilities in a vaccinated cohort**

Probability of COVID-19 infection in a vaccinated cohort

$$p(COVID|Vac) = [1 - VE_1] \times p(COVID|UnVac)$$

Probability that an COVD-19 infection requires hospitalization in a vaccinated cohort

$$p(Hosp|COVID, Vac) = [1 - VE_2^*] \times p(Hosp|COVID, UnVac)$$

Proportion of a vaccinated cohort with an COVID infection that requires hospitalization

$$p(Hosp, COVID|Vac) = p(Hosp|COVID, Vac) \times p(COVID|Vac)$$

12. Institute for Health Metrics and Evaluation (IHME). COVID-19 model update: Omicron and waning immunity. Available at: www.healthdata.org. Updated: December 22, 2021. Accessed: December 23, 2021.

13. Vaccine Effectiveness Real-time Surveillance for SARS-CoV-2 (VERSUS) Study, 5th Report. Publication date: June 6, 2022. https://www.tm.nagasaki-u.ac.jp/versus/results/20220608.html. Accessed May 17, 2023.

14. Vaccine Effectiveness Real-time Surveillance for SARS-CoV-2 (VERSUS) Study, 8th Report. Publication date: February 2, 2023. https://www.tm.nagasaki-u.ac.jp/versus/results/20230202.html. Accessed August 21, 2023.

15. Vaccine Effectiveness Real-time Surveillance for SARS-CoV-2 (VERSUS) Study, 7th Report. Publication date: December 12, 2022. . https://www.tm.nagasaki-u.ac.jp/versus/results/20221221.html. Accessed May 17, 2023.

16. Higdon MM, Baidya A, Walter KK, et al. Duration of effectiveness of vaccination against COVID-19 caused by the omicron variant. *The Lancet Infectious diseases.* 2022;22(8):1114-1116.

17. Maeda H. Effectiveness of COVID-19 vaccines in Japan (VERSUS Study). Paper presented at: 97th JAID Congress2023; Toyko, Japan.
